# Disease severity-specific neutrophil signatures in blood transcriptomes stratify COVID-19 patients

**DOI:** 10.1101/2020.07.07.20148395

**Authors:** Anna C. Aschenbrenner, Maria Mouktaroudi, Benjamin Krämer, Nikolaos Antonakos, Marie Oestreich, Konstantina Gkizeli, Melanie Nuesch-Germano, Maria Saridaki, Lorenzo Bonaguro, Nico Reusch, Kevin Baßler, Sarandia Doulou, Rainer Knoll, Tal Pecht, Theodore S. Kapellos, Nikoletta Rovina, Charlotte Kröger, Miriam Herbert, Lisa Holsten, Arik Horne, Ioanna D. Gemünd, Shobhit Agrawal, Kilian Dahm, Martina van Uelft, Anna Drews, Lena Lenkeit, Niklas Bruse, Jelle Gerretsen, Jannik Gierlich, Matthias Becker, Kristian Händler, Michael Kraut, Heidi Theis, Simachew Mengiste, Elena De Domenico, Jonas Schulte-Schrepping, Lea Seep, Jan Raabe, Christoph Hoffmeister, Michael ToVinh, Verena Keitel, Gereon Rieke, Valentina Talevi, N. Ahmad Aziz, Peter Pickkers, Frank van de Veerdonk, Mihai G. Netea, Joachim L. Schultze, Matthijs Kox, Monique M.B. Breteler, Jacob Nattermann, Antonia Koutsoukou, Evangelos J. Giamarellos-Bourboulis, Thomas Ulas, German COVID-19 Omics Initiative (DeCOI)

## Abstract

The SARS-CoV-2 pandemic is currently leading to increasing numbers of COVID-19 patients all over the world. Clinical presentations range from asymptomatic, mild respiratory tract infection, to severe cases with acute respiratory distress syndrome, respiratory failure, and death. Reports on a dysregulated immune system in the severe cases calls for a better characterization and understanding of the changes in the immune system. Here, we profiled whole blood transcriptomes of 39 COVID-19 patients and 10 control donors enabling a data-driven stratification based on molecular phenotype. Neutrophil activation-associated signatures were prominently enriched in severe patient groups, which was corroborated in whole blood transcriptomes from an independent second cohort of 30 as well as in granulocyte samples from a third cohort of 11 COVID-19 patients. Comparison of COVID-19 blood transcriptomes with those of a collection of over 2,800 samples derived from 11 different viral infections, inflammatory diseases and independent control samples revealed highly specific transcriptome signatures for COVID-19. Further, stratified transcriptomes predicted patient subgroup-specific drug candidates targeting the dysregulated systemic immune response of the host.

## INTRODUCTION

Pandemic spread of the recently emerged coronavirus, severe acute respiratory syndrome-coronavirus 2 (SARS-CoV-2), has resulted in over 9.2 million confirmed infected individuals and over 470,000 deaths worldwide (WHO, covid19.who.int, as of June 24^th^, 2020) from the resulting severe respiratory illness, called coronavirus disease 2019 (COVID-19) *(1–3)*. Based on clinical observations, it has become clear that there is great variety in disease manifestation, ranging from asymptomatic cases, to flu-like symptoms, to severe cases needing mechanical ventilation, to those who do not survive *(4–8)*. Increasing evidence suggests that the immune system plays a pivotal role in determining the severity of the disease course and it has been suggested that different molecular phenotypes might be responsible for the heterogenous outcome of COVID-19 *(9–11)*. Identifying these molecular phenotypes might not only be important for a better understanding of the pathophysiology of the disease, but also to better define patient subgroups that are more likely to benefit from specific therapies *(12–17)*. Indeed, while vaccines are still under development, finding an effective and patient-tailored therapeutic management for COVID-19 patients including targeting derailed immune mechanisms *(18, 19)* is key to mitigate the clinical burden as well as to prevent further disease fatalities *(15, 16)*.

The analysis of peripheral blood-derived immune parameters in inflammatory and infectious diseases either by classical testing, including flow cytometry and serum protein measurements, or omics technologies, including transcriptomics, has been proven very valuable in the past *(20–28)*. In COVID-19 patients, monitoring peripheral blood as a proxy for the ongoing changes within the circulating cells of the immune system, has revealed lymphopenia to correlate with disease severity *(29)*. Similarly to SARS-CoV and MERS-CoV infections, hyperinflammation due to excessive release of proinflammatory cytokines is often observed in severe COVID-19 patients as increased serum IL-6 levels correlate with respiratory failure and adverse clinical outcomes *(9, 30, 31)*.

While one can envision mild and/or early cases to benefit from antiviral drug treatments currently under clinical investigation, more severe cases may benefit from treatment to mitigate the excessive systemic immune reactions resulting in progressing pneumonia and even respiratory failure associated with severe COVID-19 *(4–9)*. The detrimental role of the systemic inflammation in the late phase of the disease has become clear, as the cytokine storm has been associated with disease morbidity *(6, 9, 30–33)*. Thus, a better understanding of the dysregulation of the host response to the infection leading to immunopathology is urgently needed to dissect and comprehend the immune parameters accompanying the heterogeneous disease severity seen upon SARS-CoV-2 infection.

Based on previous experience with other infectious diseases *(20–26)*, we hypothesized that whole blood transcriptomes should allow us to 1) determine immune cellular characteristics and functions in COVID-19 patients, 2) reveal heterogeneous molecular phenotypes of patients with similar clinical presentation, 3) define commonalities and differences of COVID-19 in comparison to other inflammatory conditions and 4) predict potential drug repurposing that might counteract observed immune dysregulations.

Here, by using blood transcriptomes, we provide evidence for molecular subtypes within the immune response of COVID-19 patients beyond distinguishing mild and severe cases only. In addition, molecular changes in blood of severely affected patients are strikingly associated with changes in the granulocyte compartment. Furthermore, blood transcriptomes of molecular subtypes of COVID-19 patients seem to be unique in comparison to more than 2,600 samples derived from other infections, inflammatory conditions and controls. Finally, by reverse drug target prediction using patients’ blood transcriptomes revealed known as well as additional new potential targets for further evaluation. Our data might also serve as a starting point for a large-scale assembly of molecular data collected during currently ongoing and future therapy trials for COVID-19 patients based on whole blood transcriptomes.

## RESULTS

### Whole blood transcriptomes reveal diversity of COVID-19 patients not explained by disease severity

To investigate the host immune response of COVID-19 patients in a systematic approach, whole blood transcriptomes were analyzed from 39 patients and 10 control donors recruited at the same hospital by RNA-sequencing (RNA-seq, **Fig. 1a**). Two-dimensional data representation using principal component analysis (PCA) showed separation of COVID-19 and control samples (**Fig. 1b**). Differential expression analysis identified 2,289 upregulated and 912 downregulated genes comparing COVID-19 and control samples (FC>|2|, padj<0.05 / **Fig. 1c**). Upregulated genes showed greater fold changes than the downregulated genes (**Fig. 1d**). Of note, *CD177*, markedly expressed in neutrophils *(34, 35)*, was the most prominently upregulated gene with the lowest p-value. Heightened expression was further found for several granulocyte- and monocyte-associated molecules, such as Eosinophil-derived neurotoxin *(RNASE2)*, Haptoglobin *(HP)*, Neutrophil elastase *(ELANE)*, Olfactomedin 4 *(OLFM4)*, Myeloperoxidase *(MPO)*, Resistin *(RETN)*, matrix metalloproteinases *(MMP8, MMP9)*, and alarmins *(S100A8, S100A9, S100A12)*, as well as for cell cycle progression-associated genes *(G0S2, CDC6, CDC25A)*, type I interferon (IFN)-induced genes *(IFI27, IFITM3*, CD169 *(SIGLEC1)*), but also genes with immunosuppressive functions *(IL10, SOCS3*, Arginase *(ARG1)*). Downregulated genes included many lymphocyte-associated factors, such as *NELL2, RORC, KLRB1*, TCF1 *(TCF7)*, Calcipressin-3 *(RCAN3), BACH2*, or *LEF1* (**Fig. 1d, Table S1**). Functional analysis of the differentially expressed genes (DEGs) by gene ontology enrichment analysis (GOEA) revealed granulocyte and complement activation-associated terms enriched in the upregulated DEGs and lymphocyte differentiation and T cell activation for the downregulated DEGs (**Fig. 1e**).Interestingly, the T cell activation-associated genes accounting for the enrichment of this term for the upregulated DEGs included *IL10* and *CD274* (PD-L1) pointing at suppressive T cell functionality (**Table S1**).

**Figure 1.**
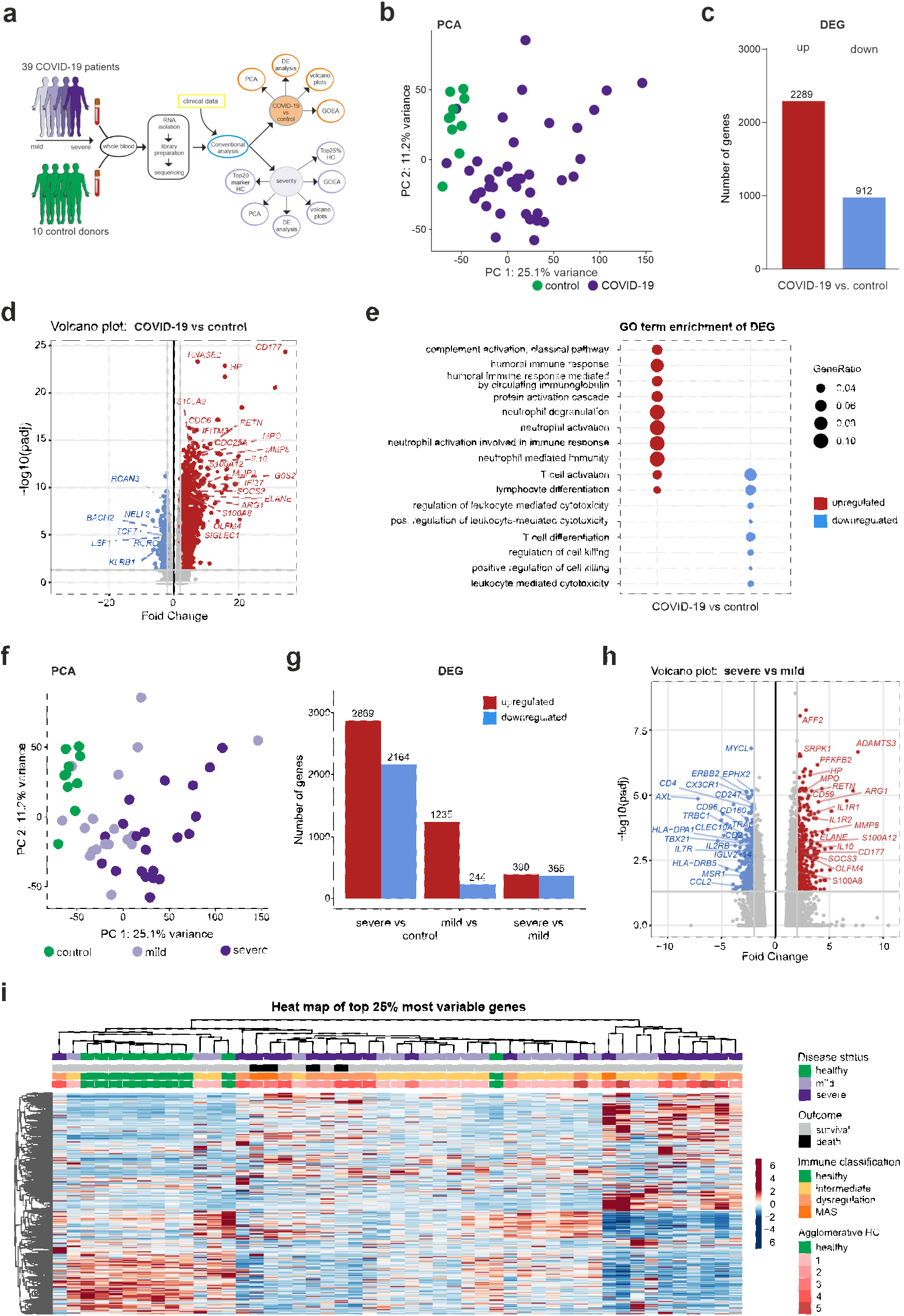
Whole blood transcriptomes reveal diversity of COVID-19 patients not explained by disease severity. **(a)** Schematic workflow for analysis of whole blood transcriptome data. **(b)** PCA plot depicting relationship of all samples based on dynamic gene expression of all genes comparing COVID-19 and control samples. **(c)** Number of significantly upregulated (red) and downregulated (blue) genes (FC>|2|, FDR-adj. p-value<0.05) comparing COVID-19 and control samples. **(d)** Volcano plot depicting fold changes (FC) and FDR-adjusted p-values comparing COVID-19 and control samples. Differentially expressed up- (red) and downregulated genes (blue) are shown and selected genes are highlighted. **(e)** Plot of top 10 most enriched GO terms for significantly up- and downregulated genes, showing ratio of significantly regulated genes within enriched GO terms (GeneRatio). **(f)** PCA plot depicting relationship of all samples based on dynamic gene expression of all genes comparing mild and severe COVID-19 as well as control samples. **(g)** Number of significantly upregulated (red) and downregulated (blue) genes (FC >|2|, FDR-adj. p-value < 0.05) comparing mild and severe COVID-19 as well as control samples. **(h)** Volcano plot depicting fold changes and FDR-adjusted p-values comparing mild and severe COVID-19 as well as control samples. Differentially expressed up- (red) and downregulated genes (blue) are shown and selected genes are highlighted. **(i)** Hierarchical clustering map of 25% most variable genes between control patients, COVID-19 mild or severe patients, with additional annotation of disease outcome, hierarchical agglomerative clustering of clinical parameters COVID-19, and the groups defined by agglomerative clustering.

In the current cohort, 51% of COVID-19 patients required intubation (**Table S2**). Given the heterogeneous nature of clinical manifestation of COVID-19, we sought to stratify the transcriptomic profiles by disease severity based on intubation status. Indeed, samples from patients with mild disease (requiring no intubation) clustered more closely to the control samples, while those of severe cases scattered away in the PCA (**Fig. 1f**). Consequently, there was a greater number of DEGs in blood samples from severe COVID-19 patients than in mild patients when compared to controls (**Fig. 1g**). Many of the DEGs found in the COVID-19 vs control comparison (**Fig. 1d**) were also found when separating the COVID-19 samples by severity (**Fig S1a,b**). Both, severe and mild COVID-19 in comparison to controls sh The SARS-CoV-2 pandemic is currently leading to increasing numbers of COVID-19 patients all over the world. Clinical presentations range from asymptomatic, mild respiratory tract infection, to severe cases with acute respiratory distress syndrome, respiratory failure, and death. Reports on a dysregulated immune system in the severe cases calls for a better characterization and understanding of the changes in the immune system. Here, we profiled whole blood transcriptomes of 39 COVID-19 patients and 10 control donors enabling a data-driven stratification based on molecular phenotype. Neutrophil activation-associated signatures were prominently enriched in severe patient groups, which was corroborated in whole blood transcriptomes from an independent second cohort of 30 as well as in granulocyte samples from a third cohort of 11 COVID-19 patients. Comparison of COVID-19 blood transcriptomes with those of a collection of over 2,600 samples derived from 11 different viral infections, inflammatory diseases and independent control samples revealed highly specific transcriptome signatures for COVID-19. Further, stratified transcriptomes predicted patient subgroup-specific drug candidates targeting the dysregulated systemic immune response of the host.ared neutrophil-specific *CD177* and *HP* expression among the most upregulated DEGs, as well as lymphocyte-associated genes such as ABLIM1, NELL2, RCAN3, RORC, KLRB1, among the downregulated genes (**Fig. S1a,b**). GOEA reflected these findings (**Fig. S1c**). Although all samples from COVID-19 patients showed functional enrichment for granulocyte/neutrophil activation-associated terms in general, direct comparison of severe and mild COVID-19 patients revealed this to be a heightened characteristic of the immunoprofiles in severe COVID-19 (**Fig. S1c**). Upregulated DEGs in the severe vs. mild sample comparison included *CD177*, Haptoglobin *(HP)*, Neutrophil elastase *(ELANE)*, Olfactomedin 4 *(OLFM4)*, Myeloperoxidase *(MPO)*, Resistin *(RETN)*, matrix metalloproteinase *MMP8*, and alarmins *(S100A8, S100A12)*. Whereas the type I IFN-response genes, such as *IFI27* or *IFITM3*, were not differentially regulated in severe vs. mild samples, expression of immunosuppression-associated factors was more pronounced in severe COVID-19 patients *(IL10, SOCS3*, Arginase *(ARG1)*) (**Fig. 1h, Table S1**). Moreover, blood transcriptomes from severe cases showed decreased expression of lymphocyte-associated genes, such as the T cell receptor chains *(TRAC, TRBC1)*, CD3 zeta chain *(CD247), CD4, CD2,IL2RB*, TBET *(TBX21), IL7R*, as well as monocyte-associated genes, such as MHC class II molecules *(HLA-DPA1, HLA-DRB5)*, fractalkine receptor *(CX3CR1)*, Macrophage scavenger receptor *(MSR1)*, or *CCL2* (**Fig. 1h, Table S1**). Differences in gene expression were not restricted to granulocyte and T cell functions only: assessing the changes in defined gene groups, e.g. transcription factors, epigenetic regulators, surface or secreted molecules, we observed many significant changes in genes that are not restricted to granulocytes or T cells, clearly indicating that other cell types are also transcriptionally altered in COVID-19 patients (**Fig. S1d**).

Distribution of the COVID-19 samples in the PCA revealed heterogeneity in the transcriptomic profiles (**Fig. 1f**), which might be due to clinical heterogeneity (**Table S2**). In order to investigate this further, the top 25% of the most variable expressed genes were visualized in a heat map and samples sorted by unbiased hierarchical clustering based on their transcriptomic profiles, which resulted in more than three clusters suggesting higher transcriptional heterogeneity as explained by mild and severe COVID-19 cases vs control (**Fig. 1i**). Strikingly, neither disease, disease severity, nor the inclusion of outcome or immune classification *(31)*, sufficiently explained the structure in the data. In order to get a better clinical understanding of the transcriptional data, we included further clinical parameters and grouped the COVID-19 patients accordingly (**Fig. 1i**). We therefore performed agglomerative hierarchical clustering using the clinical parameters that contributed most to the transcriptional differences observed across the first principal component of the dataset (r-adjusted square ≥0.1, **Fig. S1e**). The COVID-19 patients were clustered into five clinical groups, which was the optimal number of clusters at which the intra-group variance was low and the ‘clusters distance’ remained high (**Fig. S1f,g**). However, comparison of this clinical parameter-based grouping of the COVID-19 patients did not match the transcriptional variability observed in the data either (**Fig. 1i**), arguing that additional molecular parameters must exist that better define the blood transcriptome structure and thereby more accurately dissect heterogeneity of the clinical manifestation of COVID-19.

### Co-expression analysis discloses COVID-19 subgroups with distinct molecular signatures

Classical approaches to analyze the transcriptome data by using differential gene expression analysis based on sample groups defined by a selection of clinical parameters precluded dissection of the heterogeneity of the host immune response towards SARS-CoV-2 infection, which is evident in the high-parameter space of the transcriptome (**Fig. 1**). Co-expression analysis on the other hand identifies similarly regulated genes across samples, groups these genes into modules, which can then be explored for each patient sample individually or for entire patient groups. Applying such an approach using our established CoCena^2^ pipeline [https://github.com/Ulas-lab/CoCena2] (**Fig. 2a**) for all 49 samples (39 COVID-19, 10 control) independent of their clinical annotation disclosed 10 co-expression modules, designated by color indianred to darkgrey, across a total of 6,085 genes included in the analysis (**Fig. S2a**). Hierarchical clustering of the samples based on their group fold changes (GFCs) for each module revealed a data-driven patient stratification assorting the samples into six groups (**Fig. S2b**), which were subsequently used in all following analyses: five different COVID-19 sample-containing groups, which only partially grouped by disease severity and illustrated heterogeneity of the immune response in COVID-19 patients, plus one group containing all control as well as four COVID-19 samples (**Fig. 2b**+**S2c**). Overlaying this information onto the original PCA reflected structured sample stratification as the newly defined groups clustered together (**Fig. S2d**). GFC analysis of the newly generated groups revealed group-specific enrichment of co-expressed gene modules (**Fig. 2c**). GOEA on each of the modules identified associated gene signatures displaying distinct functional characteristics, which distinguish the different sample groups G1-G6 (**Fig. 2d+S3, Table S3**). For example, ‘inflammatory response’ was enriched in modules maroon, lightgreen, pink, and darkgrey, all characteristic for sample groups G1 and G2 to different extents, indicating these to possibly undergoing a more vigorous inflammatory immune reaction (**Fig. 2c+d**). Of note, G1 and G2 harbour a great fraction of samples from patients with severe COVID-19 (**Fig. 2b**). Only a slight increase in the inflammation-associated module maroon, an increase in expression in the genes of darkorange (enriched in oxidative phosphorylation, mTORC1 signaling and cell cycle-associated genes), as well as a loss of expression in the gold module (connected to estrogen response genes and IL2 signaling) was indicative of the G4 sample group. G6, encompassing all control samples, was not associated with any modules connected to inflammatory processes, but showed higher expression of indianred, steelblue and gold, all functionally enriched basic cellular and metabolic processes. Extended analysis of the lightgreen module, containing 987 genes, revealed a prominent enrichment of granulocyte/neutrophil activation-related signatures (**Fig. 2e, Table S3**). To further explore this neutrophil activation signature association, we investigated possible co-expression patterns of long non-coding RNAs (lncRNA) that were reported as regulators of granulocyte function *(36)*. *CYTOR* (also known as *Morrbid)* is a lncRNA that mediates survival of neutrophils, eosinophils, and classical monocytes in response to pro-survival cytokines *(36)*, and interacts with the protein-coding RNAs for the catalytic PI3K isoform Phosphatidylinositol-4,5-bisphosphate 3-kinase catalytic subunit beta *(PIK3CB)* and the filament Vimentin *(VIM) (37)*. Interestingly, expression of *CYTOR* was significantly increased in severe COVID-19 patient group G1 (p<0.001) and correlated with both *PIK3CB* (r= 0.53, p<0.001) and *VIM* (r= 0.55, p<0.001) (**Fig. 2f**).

**Figure 2.**
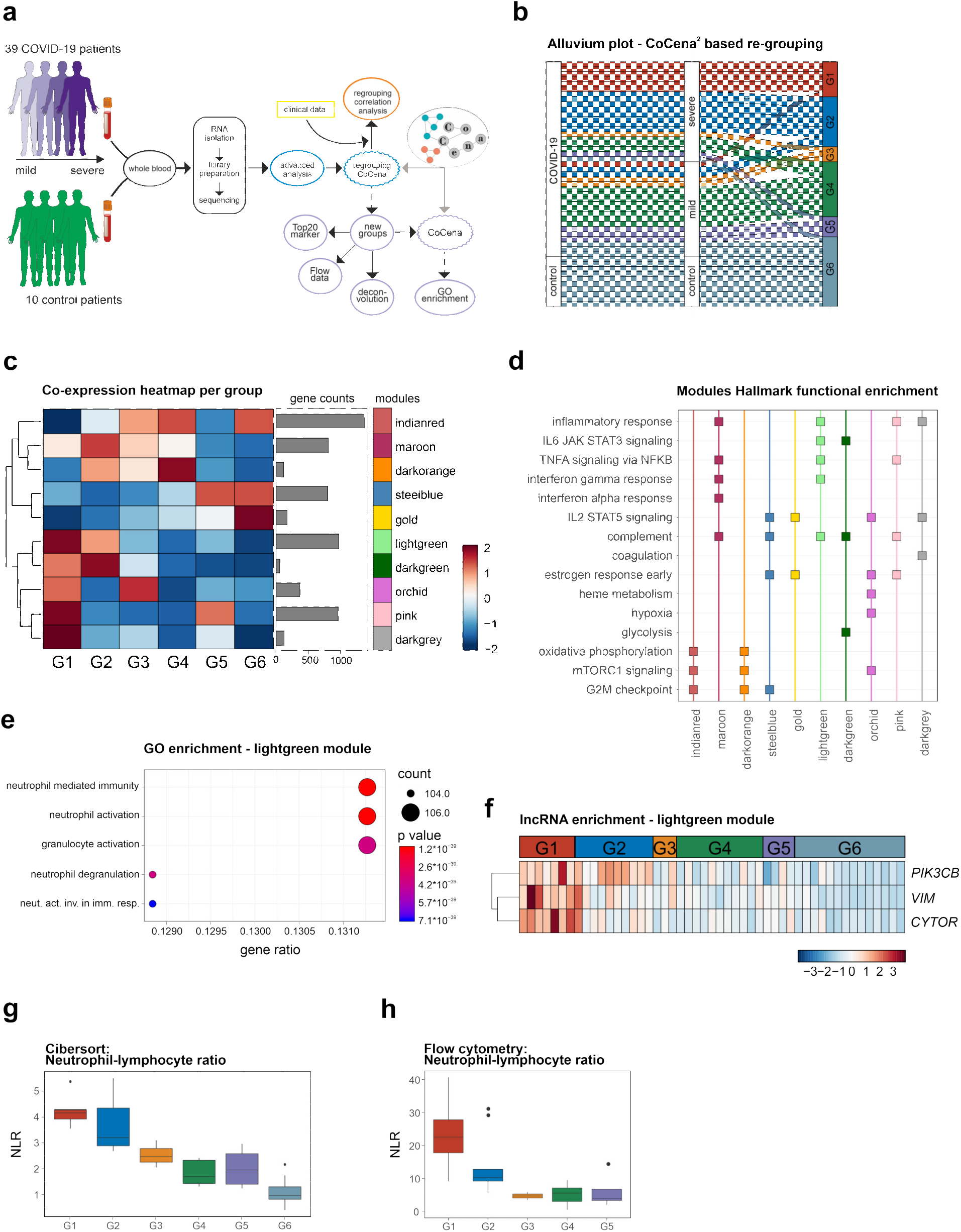
Co-expression analysis discloses COVID-19 subgroups with distinct molecular signatures. **(a)** Schematic overview of the analysis performed on the whole blood samples. **(b)** Alluvium plot visualizing the distribution of the samples according to different grouping; disease status, severity and data-driven sample groups. **(c)** Group fold change heat map and hierarchical clustering for the six data-driven sample groups and the gene modules identified byCoCena^2^ analysis. **(d)** Functional enrichment of CoCena^2^-derived modules using the Hallmark gene set database. Selected top terms were visualized. **(e)** Functional enrichment of CoCena^2^ module lightgreen using GO gene set database. Top 5 terms were visualized. **(f)** Heat map presenting the normalized expression values of the lncRNA *CYTOR*, and protein coding RNAs *PIK3CB* and *VIM* from the lightgreen CoCena^2^ module. **(g)** Neutrophil-lymphocyte ratio plot after cell type deconvolution at lineage level. **(h)** Neutrophil-lymphocyte ratio across the six data-driven sample groups. Box plots show median with variance, with lower and upper hinges representing the 25^th^ and 75^th^ percentile, respectively.

Next, we asked whether the enrichment for neutrophil activation-associated signatures in G1 and G2 is attributed to an increased relative number of granulocytes within the whole blood sample. Deconvolution of the expression values using linear support vector regression *(38)* showed increased relative percentages of neutrophils especially in G1 and G2 (**Fig. S2e**). G5, on the other hand, clearly displayed an increased percentage of monocytes. At the same time, lymphocyte enrichment was found to be reduced in the COVID-19 sample groups, most prominently in G1 and G2 (**Fig. S2e**). The linear deconvolution results were then validated by flow cytometry. Blood composition of COVID-19 donors confirmed an increased number of neutrophils and a decreased number of lymphocytes especially in G1 and G2 (**Fig. S2f**). As a result, the neutrophil-lymphocyte ratio (NLR), a clinical marker proposed for disease severity as it has been associated with an increased systemic inflammation *(39, 40)*, was markedly elevated in G1 and G2 compared to the control sample-containing G6, both in the computationally deconvoluted results (**Fig. 2g**) as well as measured by flow cytometry (**Fig. 2h**). Interestingly, in context of the observation that men more often progress to severe COVID-19 than women *(41)*, G1 encompasses samples from solely male patients (**Fig. S2c**). Analysis of the top 20 differentially expressed transcription factors, epigenetic regulators, surface or secreted proteins for the six sample groups confirmed an increased inflammatory state, again most remarkably for G1 and G2, evident from the transcription factors of the STAT family, *STAT1, STAT3, STAT5B* and *STAT6*, surface marker *CSF3R* (G-CSF) or *FCGR3B* (CD16b), the secreted factors *GRN* or *IL1B*, or the epigenetic regulator *PADI4* (PAD4) (**Fig. S2h**).

We confirmed our findings of distinct molecular phenotypes in the blood of COVID-19 patients in a second independent cohort. Thirty patients, severely affected by SARS-CoV-2 infection, were sampled upon admission to the ICU. We stratified the obtained blood transcriptomes based on the module signatures from the co-expression analysis (**Fig. 2c**). The samples of the second cohort were filtered for the genes present in the COVID-19 co-expression network, group fold changes were calculated across all patients individually, and sample groups G1-G6 assigned according to their combinatorial module expression (**Fig. S4a**). Controls from the first cohort were included for comparison. Interestingly, in these ICU patients, we noted the transcriptome profiles from the second cohort to show greatest similarity to G1 and G2, which is in line with their severe phenotypes and our findings from the first cohort. Hierarchical clustering of the samples based on their group fold changes for each module stratified the samples of the second cohort into four groups (**Fig. S4b**). The control samples from the first cohort built one separate group, which we designated again as G6. To allow for group-specific comparison to the stratification within the first cohort (**Fig. 2c**), we calculated the mean GFCs of the four groups identified in the second cohort (**Fig. S4c**). Second cohort samples of the first group showed enrichment in modules lightgreen, pink and darkgrey and were thus assigned most similar to G1; the third group of the new samples showed enrichment in modules maroon and darkorange, most similar to G2; and the remaining samples were stratified into an intermediate group exhibiting stronger expression of genes from the darkorange as well as pink module indicating characteristics of both G1 and G2 (**Fig. S4c**).

Collectively, co-expression analysis (CoCena^2^) in whole blood transcriptomes reveals at least five molecular phenotypes of the host’s immune response in COVID-19 patients with at least two different groups in clinically described severe COVID-19 patients. The two molecularly defined groups G1 and G2 are transcriptionally characterized by a pronounced neutrophilic signature, at the same time distinct in other cellular characteristics. Such molecular classification might serve as a basis for identifying clinical surrogates for patient stratification. Since whole blood transcriptomics captures functional changes in the host’s peripheral immune response across all cell types, we next sought a more detailed investigation of the granulocyte compartment within the framework of the newly identified subgroups.

### Granulocytes from severe COVID-19 patients show a simultaneous increase in inflammatory and suppressive signatures

To investigate whether the activation signatures seen in whole blood of COVID-19 patients are not only due to disease-associated increase of the neutrophil population, granulocytes were sequenced and transcriptomes were analyzed from 11 longitudinally sampled patients (4 mild, 7 severe), resulting in 14 mild and 45 severe COVID-19 samples (**Fig. 3a**). Evaluation of the relative cell type composition within each sample using linear deconvolution predicted the samples to mainly consist of neutrophils, with comparable fractions of 78.8% on average (**Fig S5a**). Exploratory analysis by PCA showed a separation between mild and severe COVID-19 patients’ granulocyte samples, especially for the day 1-14 groups (**Fig. 3B**). Differential expression analysis identified 1,496 upregulated and 1,440 downregulated genes comparing severe and mild samples from day 1-14 after first symptoms, while comparison at a late disease stage showed less differences on gene level (380 up-, 307 downregulated genes / FC>|2|, padj<0.05 / **Fig. 3c, Table S4**). Whole blood transcriptome analysis showed enrichment of neutrophil activation-associated signatures (**Fig. 2**). Excluding the bias of alterations in neutrophil population size across conditions, gene set enrichment analysis on granulocyte samples now uncovered that differentially expressed genes between severe and mild COVID-19 patients are indeed characterized by an increase in granulocyte activation-associated factors (**Fig. S5b**). *CD177* is part of the granulocyte activation gene set and was indeed markedly increased in severe (day 1-14) compared to mild (day 1-14) COVID-19 samples (**Fig. 3d**). Also, the alarmin *S100A6* exhibited heightened expression in granulocytes from severe COVID-19 patients (**Fig. 3d**).

**Figure 3.**
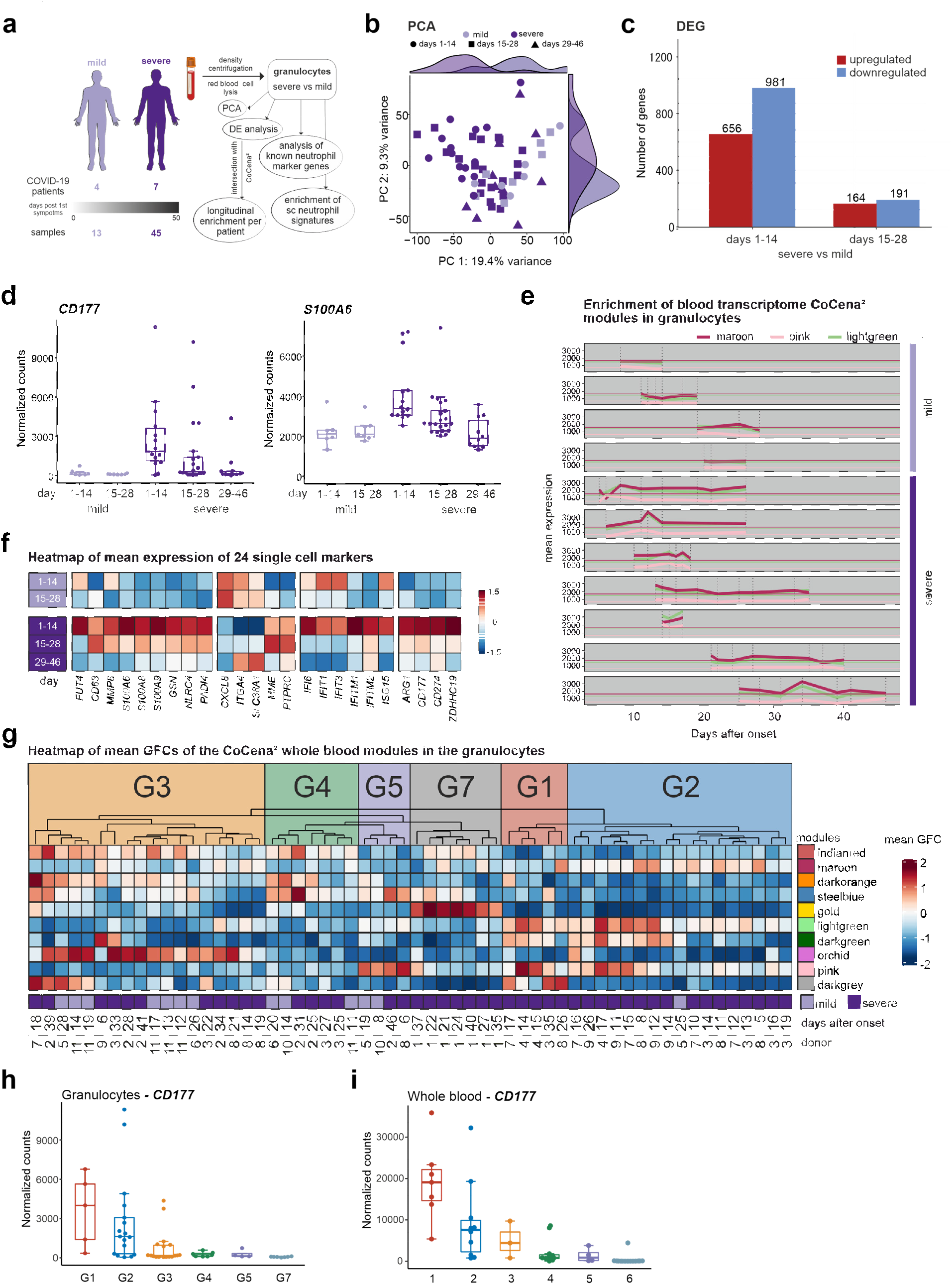
Granulocytes from severe COVID-19 patients show a simultaneous increase in inflammatory and suppressive signatures. **(a)** Schema of sample processing and analysis. **(b)** PCA of all genes within the dataset mapped by COVID-19 severity status. **(c)** Bar plot of DEGs between severe and mild COVID-19 patients at day 1-14 (left) and day 15-28 (right) (FC>|2|, FDR-adj. p-value <0.05). **(d)** Boxplot of *CD177* (left) and *S100A6* (right) in mild and severe COVID-19 patients at day 1-14 and 15-28. **(e)** Mean of group fold changes (GFCs) of the modules maroon, pink and lightgreen in the granulocyte samples over time. Patients are grouped according to severity mild (top) and severe (bottom). Samples are ordered by the days after disease onset. Maroon, pink and lightgreen lines represent the overall mean of GFCs in the mild patient group. **(f)** Heat map of mean expression of 24 markers in mild (top) and severe (bottom) patients ordered by days after disease onset bins (day 1-14, 15-28 and 29-46). **(g)** Heat map of mean GFCs of the CoCena^2^ whole blood modules in the granulocyte samples from each individual patient. Patients are clusters by the mean GFC module expression. Severity patterns found in the whole blood CoCena^2^ network were identified and patients groups were assigned accordingly (G1-G6). Patients with a distinct GFC expression pattern were labeled as G7. **(h)** Box plot of *CD177* expression in granulocytes grouped by G1-G7. **(i)** Box plot of *CD177* expression in whole blood grouped by G1-G6.

Next, we used the CoCena^2^ modules from the whole blood analysis (**Fig. 2c**) to identify modules that are actually driven by alterations in neutrophil activation instead of a mere increase in the neutrophil population. The genes from the modules were filtered by the union of the upregulated genes between severe and mild COVID-19 patients (either comparing at day 1-14 or 15-28 / **Fig. S5c**). After filtering, the number of DEG between granulocyte samples from mild and severe COVID-19 exceeded 100 genes per module in three of the modules. Among those modules, the genes identified in whole blood transcriptomes within the lightgreen module showed the highest overlap of 45%, maroon of 29% and pink of 7% with genes upregulated in granulocytes of severe COVID-19 patients. We then investigated the expression pattern of those modules for each individual patient in a longitudinal fashion (**Fig. 3e**). In concordance with the whole blood CoCena^2^ results (**Fig. 2c**), modules lightgreen, maroon and pink showed a continuously elevated mean expression in the severe compared to the mild COVID-19 patients, indicated by horizontal lines showing the mean expression of the respective modules calculated only for the mild patients (**Fig. 3e**). This effect was observed irrespective of donor and of days after symptoms onset.

Recently, heterogeneity of neutrophils with distinct subsets associated with disease severity and phase was revealed by single cell RNA-seq analysis in blood of COVID-19 patients *(42)*. Enrichment of the three signatures that related to severe COVID-19, in our granulocyte samples demonstrated that the findings obtained in the single-cell study were also discernible in bulk data and the results in accordance to the reported phenotypes: premature/immature, severe inflammatory, as well as severe suppressive subset marker genes were markedly enriched in granulocytes from severe COVID-19 patients in the present study (**Fig. S5d**). Further analysis of this observation on the gene level displayed the heightened expression of pre-/immature neutrophil-associated markers in severe COVID-19 patients’ granulocytes, such as CD15 *(FUT4)*, metalloproteinase *MMP8*, alarmins *(S100A8/9)*, NET formation-involved *PADI4*, or *NLRC4*, for which activating mutations have been reported to overtly trigger the inflammasome and thereby increase the risk to develop autoinflammatory syndrome *(43, 44)* (**Fig. 3f**). Marker genes attributed to the “mild mature activated” neutrophil subset *(42)*, such as *ITGA4*, or *SLC38A1*, were indeed elevated as well in the mild COVID-19 patients’ granulocytes of this study. In line with the single cell study, signs of an interferon response were observed irrespective of disease severity *(IFIT1, IFIT3, ISG15)*, while only severe COVID-19 patients’ granulocytes featured expression of genes with suppressive functionality, such as *ARG1* or PD-L1 *(CD274)* (**Fig. 3f**),

We next stratified the granulocyte samples based on the module signatures from the whole blood analysis. The granulocyte samples were filtered for the genes present in the COVID-19 co-expression network (**Fig. 2c**) and the group fold changes were calculated across all patients individually, sample groups G1-G6 were assigned according to their combinatorial module expression (**Fig. 2c+3g**). For example, samples attributed to G1 showed high enrichment scores in modules lightgreen, darkgreen and pink, whereas those assigned as G2 additionally expressed the maroon module. Samples with the indianred/darkorange combination were designated as G4. Assessment of the combinatorial enrichment scores for the different modules did not lead to a corresponding sample group for all longitudinal samples from patient 1, hence it was assigned as G7. Re-analysis of *CD177, NLRC4, ARG1*, and PD-L1 *(CD274)* as a function of the assigned sample groups (**Fig. 3b-d**), showed increased expression in G1 and G2 in relation to the other groups (**Fig. 3h+S5e**). Interestingly, the stratified patient groups in the whole blood data also depicted increased expression in G1 and G2 in comparison to the control-containing G6 (**Fig. 3i+S5f**).

Analysis of granulocyte samples from COVID-19 patients proved that, in addition to the relative increase in neutrophils in severe COVID-19 cases, there are indeed alterations in the transcriptional program of these cells themselves. We found enrichment of signatures typical of pre-/immature neutrophils and evidence of simultaneous inflammatory and suppressive features, arguing for a dysregulation in the peripheral granulocyte compartment. Importantly, transferring these findings back to the whole blood analysis showed that the granulocyte phenotypes were still observable within the whole blood transcriptomes.

### Integration with signatures from other diseases reveals COVID-19-specific characteristics

Putting COVID-19 into context of other known diseases, we compiled whole blood transcriptomes from 11 further diseases, including several viral and bacterial infections as well as immune-related disorders into one large dataset encompassing a total of 2,817 samples including the 39 COVID-19 samples from this study (**Fig. 4a, S6a, Table S5**). All in all, the dataset contains three other viral infection studies (Chikungunya *(26)*, HIV *(23)* and Zika *(45)*, n=466), seven bacterial infection studies (tuberculosis *(20–23, 46)*, bacterial sepsis and systemic inflammatory response syndrome (SIRS, n=1,578) *(24)*, six inflammatory/autoimmune studies (systemic lupus erythematosus *(47)*, Crohn’s disease, rheumatoid arthritis *(48)*, Ebola vaccination *(25)*, neonatal-onset multisystem inflammatory disease (NOMID) and macrophage activation syndrome (NLRC4-MAS) *(44)*, n=326) as well as control samples from eight different studies (n=408). To investigate how the COVID-19-specific co-expression modules can be linked to other diseases, the combined dataset was filtered for the genes present in the COVID-19 co-expression network (**Fig. 2c**) and the group fold changes were calculated across all samples (**Fig. 4b**). Additionally, cell type-specific signatures *(38)* and single cell-derived neutrophil subset signatures *(42)* (**Table S6**) were intersected with all CoCena^2^ modules. This analysis revealed that the lightgreen module shows a high (61%) neutrophil enrichment followed by module pink (38%) and maroon (32%), which is in line with a high functional enrichment for neutrophil activation in lightgreen (**Fig. 2e, Table S3**). Genes within module lightgreen were most prominently upregulated in the severe COVID-19 group (G1) as well as in sepsis and in patients with tuberculosis and HIV infection, but not in individually occurring HIV and tuberculosis (**Fig. 4b**). Enrichment of the neutrophil subset signatures revealed increased expression of genes found in pre-/immature neutrophils and those of inflammatory neutrophils associated with severe COVID-19. Many genes within module lightgreen are known to be related to induction of neutrophil extracellular traps (NET) (e.g. *PKC (49), PADI4 (50), LTB4 (51)*). Moreover, a link between excessive NET activation and tissue damage has been reported in sepsis *(52)*. Module darkgrey shares a similar expression pattern across the disease spectrum with lightgreen and contains genes involved in platelet activation. The NET–platelet– thrombin axis has been reported to be involved in the promotion of intravascular coagulation in sepsis *(53)*. The pink module shows the second highest neutrophil enrichment, which is dominated by the enrichment of pre-/immature neutrophils subtype signatures. It is strongly increased in sepsis, tuberculosis, after Ebola vaccination as well as in autoinflammatory diseases such as rheumatoid arthritis, NLRC4-MAS and NOMID, and shows slight overlap with the severe COVID-19 patients in group G1. It contains many epigenetic modifiers, such as *HDAC5, SETD1B*, or *KMT2D*, as well as *KLF2*, shown to regulate NF-κB-mediated immune functions, such as inflammation, erythropoiesis and lung development *(54)*. Maroon is the third module with predicted neutrophil enrichment, which features genes from the “severe suppressive” subset alongside the “severe inflammatory” and pre-/immature subset signatures. It is associated with COVID-19 groups G2-4 and shares this characteristic with blood transcriptomes from the response to infection with Chikungunya and Zika virus or from HIV patients suffering from tuberculosis.

**Figure 4.**
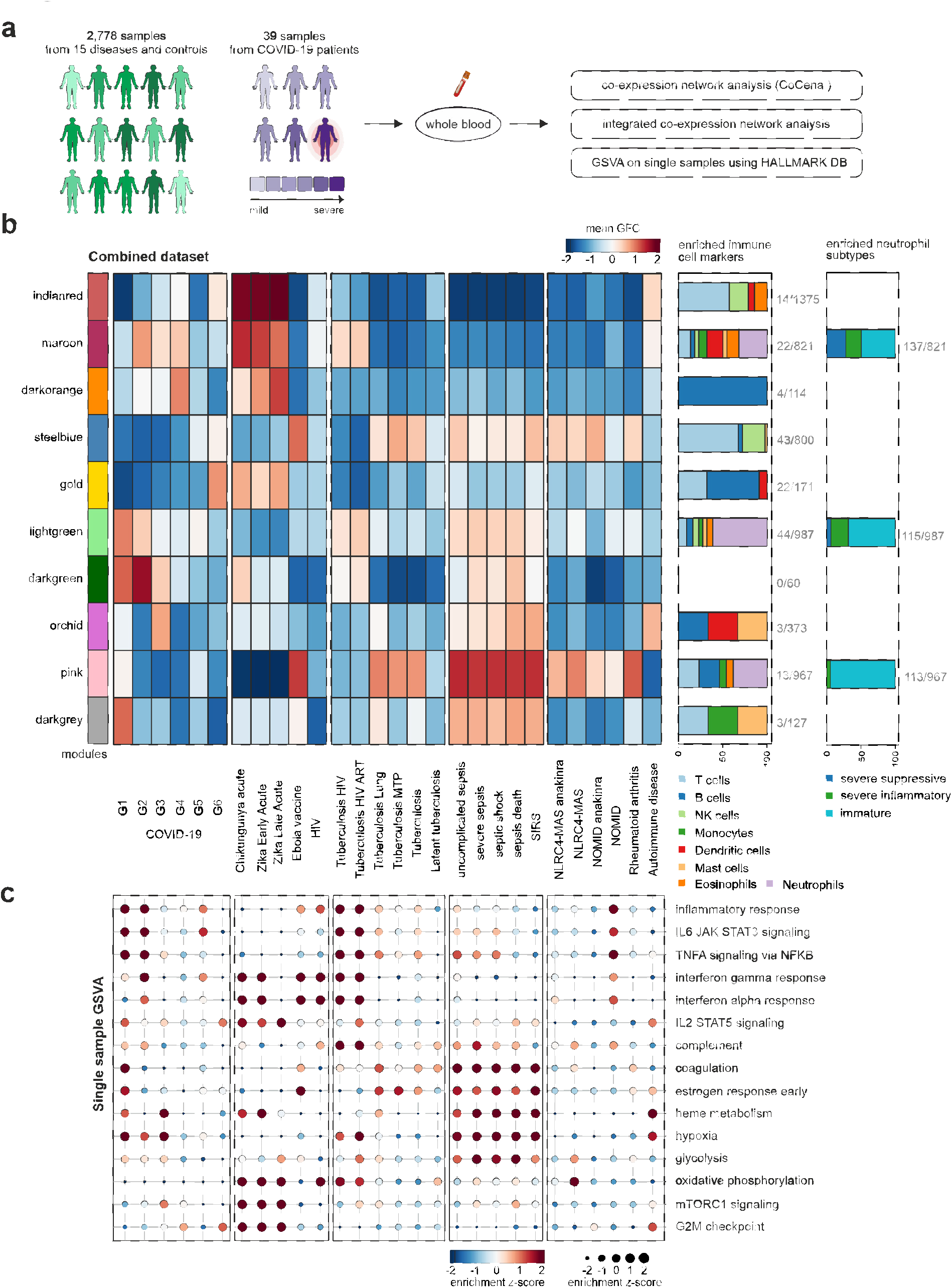
Integration with signatures from other diseases reveals COVID-19-specific characteristics. **(a)** Schema of analysis of the integrated dataset. The integrated dataset was analyzed with regard to expression patterns of the clusters previously identified in the whole blood COVID-19-specific co-expression network. **(b)** Heat map of mean group fold changes of CoCena^2^ module comparison between COVID-19 and other diseases. From left to right, the diseases are ordered by category (COVID-19, viral infections, bacterial infections and others). On the right side of the heat map, the first box plot shows the enriched immune cell markers in each module. The second box plot shows the enrichment of genes upregulated in specific neutrophil subtypes based on cross-referencing with single-cell data *(42)*. Both box plots show enriched cell types in percent of total hits, absolute hits with respect to cluster size are stated aside. **(c)** Gene set variation analysis was conducted for every single patient based on Hallmark gene sets as shown in Fig. 2D. The result was standardized to retrieve the z-scores, a disease mean was calculated and displayed as a dot plot with size and color correlating to the z-score. The labels on the x-axis are the same as in **(b)**.

A combination of single sample gene set variation analysis (ssGSVA), a non-parametric, unsupervised approach to estimate variation of gene set enrichment within each single sample, and Hallmark enrichment for each disease or inflammatory condition in the compiled dataset accentuated the findings on COVID-19 blood transcriptomes in context of the other diseases (**Fig. 4c**). ‘Interferon alpha and gamma responses’ were enriched in acute viral infections with Chikungunya and Zika virus as well as in HIV with or without concomittent tuberculosis or after Ebola vaccination, and this enrichment was shared with COVID-19 G2. ‘Inflammatory response’, ‘IL6 and TNFA signaling’ is an attribute of both, G1 and G2, to a lesser degree of G5, also Tuberculosis/HIV, and to some extent of sepsis. More prominently enriched in sepsis was ‘complement’, ‘coagulation’, ‘heme metabolism’ and ‘glycolysis’ - shared by COVID-19 G1+G3; whereas ‘oxidative phosphorylation’ and ‘mTORC1 signalling’ were seen for Chikungunya and Zika virus infections - shared to some extent with COVID-19 G3+G4.

Although we observed overlaps of gene modules enriched in COVID-19 with several other infectious and immune-related diseases, each of our molecularly defined COVID-19 patient groups was characterized by a specific combination of these modules, clearly indicating the unique biology of this SARS-CoV-2 infection-mediated immune response, which needs to be considered when developing patient-stratified therapy regimens.

### COVID-19 patient subgroup-specific signatures can be used to predict potential drug repurposing

Despite the immunologically-driven nature of COVID-19, most drugs that are currently investigated in clinical trials to combat or ameliorate COVID-19 are targeting the virus and its direct interaction partners (**Fig. 5a+S7a, Table S7**). Compounds as well as the number of clinical trials performed with anti-inflammatory, immunosuppressive, and immunomodulatory properties are immensely outnumbered by other approaches. Examining the listed target genes of currently investigated drugs in our stratified patient groups, we found 162 included in our co-expression network analysis, most of which being differentially expressed in the severe patient group G1 in comparison to G6 (**Fig. 2c+5b**). In addition, many of the regulated genes in our patient signatures are clearly not affected by the drugs that are currently investigated against COVID-19. The immunopathologies seen in COVID-19 patients, especially past their second week of symptoms, demand a host-directed, immune system-focused therapy.

**Figure 5.**
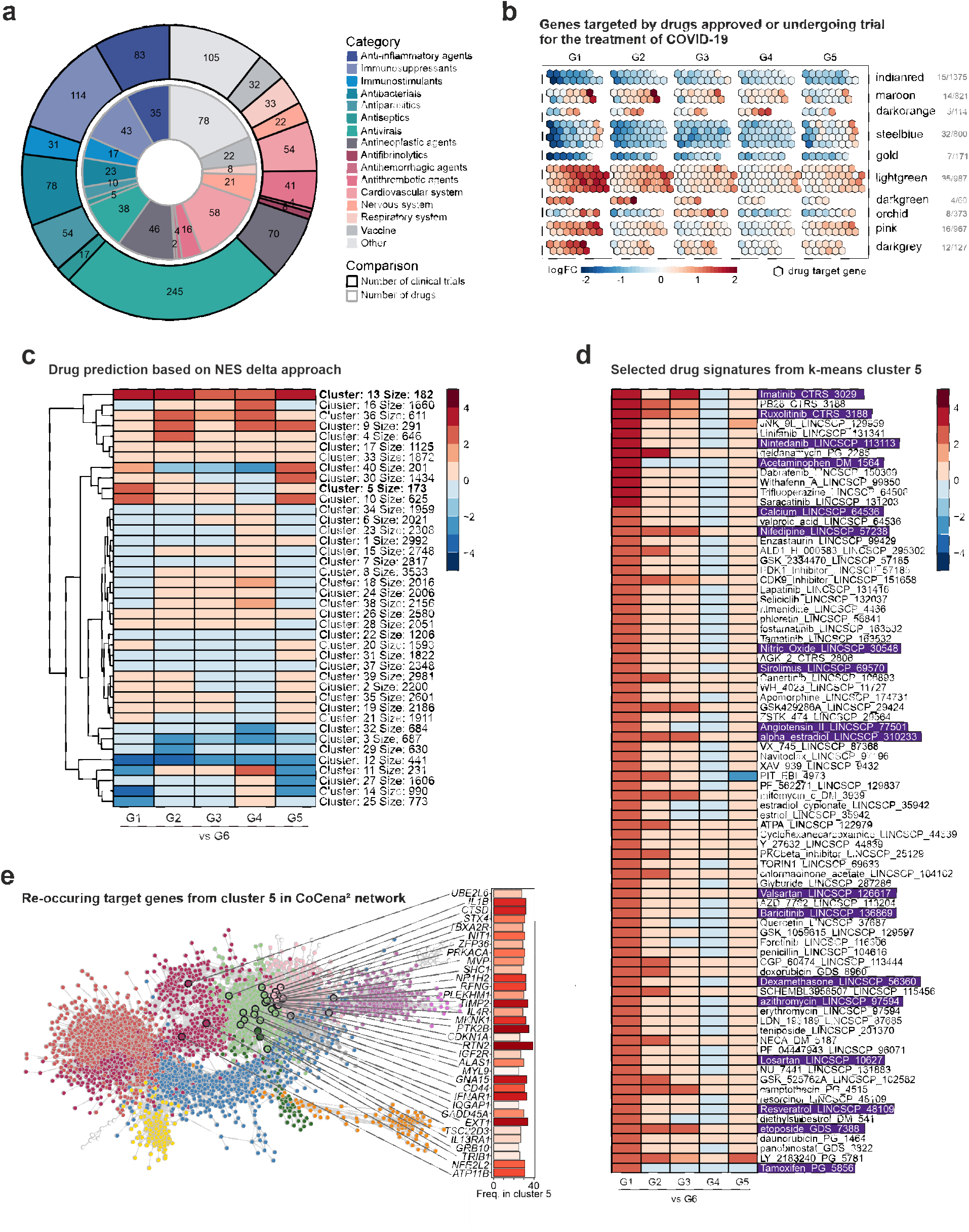
Patient subgroup-specific signatures can be used to predict potential drug targets. **(a)** Overview of drugs currently used, investigated or recommended for the treatment of COVID-19 patients. The inner circle represents the number of drugs for the representative drug categories, the outer circle represents the number of clinical trials of drugs for the respective drug categories. **(b)** Visualization of genes targeted by drugs approved or undergoing trial for the treatment of COVID-19 patients included in the whole blood co-expression network. Numbers of such genes from each module are designated on the right of the panel. Genes are represented as hexagons and colored by the expression fold change between COVID-19 patient severity group (G1-G5) and the control group (G6) (upregulated: red, downregulated: blue, not regulated: grey). **(c)** Drug predictions based on ΔNES score of drug signatures in regard to diseased patient group-specific gene expression patterns (G1-5 vs G6). Signatures were clustered by k-means clustering. A high ΔNES score accounts for drug signatures which counteract the gene expression of the patient group they are compared to. Drug signatures with a negative ΔNES score induce a gene expression pattern similar to the input. The number of signatures within a cluster determines its size. **(d)** Display of selected drug signatures from k-means cluster 5 from **(C)** showing the highest ΔNES score in the most severe COVID-19 patient group G1 and the least effect in patient group G4. **(e)** Visualization of recurring target genes in the G1 vs G6 comparison of cluster 5 signatures and their frequency mapped onto the CoCena^2^ network.

To identify potentially beneficial drugs, we designed an *in silico* signature-based drug repurposing approach (**Fig. S7b**). To generate input signatures of interest, we characterized our stratified sample groups by identifying differentially expressed genes between groups G1-G5 and the control group G6 (**Fig. S7c**). Most transcriptional differences were observed for G1 (up: 4,032, down: 4,729) and G2 (up: 2,336, down: 2,767), whereas group G3 (up: 1,193, down: 1,921), G5 (up: 1089, down: 1216), and especially G4 (up: 727, down: 547) were less different to G6. Only a minor fraction of 137 DEGs was shared by all 5 comparisons. The most overlap of DEGs was observed between G1 and G2, the two groups comprising mostly severe COVID-19 patients. Nevertheless, G2 was still characterized by a large number of specific DEGs (**Fig. S7c**). GOEA of the upregulated DEGs of each comparison revealed enrichment of genes in the context of ‘neutrophil activation’ and ‘coagulation’ in all comparisons (**Fig. S7d**). Humoral and B cell-mediated immunity terms on the other hand were enriched the strongest in G4-specific upregulated DEGs (**Fig. S7d**). Differential expression analysis for the stratified sample groups once more emphasized that neutrophils play a central role in the host’s immune response against SARS-CoV-2 infection. Neutrophils, as the most abundant circulating leukocytes, have become a therapeutic target of interest in multiple disease settings in recent years *(55)*. Two interesting target genes discussed in this context and already addressed in clinical trials are CXCR2 and C5AR1. Consistent with the increased NLR in G1 and G2, we observed significant upregulation of *CXCR2* and *C5AR1* in both groups (**Fig. S7e**).

Using patient cluster-specific DEGs as input (**Fig. S7c, Table S8**), we searched for compounds that evoke a reverse signature in human cells via the NIH Library of Integrated Network-Based Cellular Signatures (iLINCS) *(56)* and the Broad Institute’s Repurposing Hub *(57)*. The best counteracting signatures for each comparison were combined with signatures for all currently investigated drugs and downloaded for further analysis, resulting in about 63,000 signatures from 940 compounds/drugs. We performed gene set enrichment analysis for all signatures against our COVID-19 patient comparisons and calculated the difference of the up- and downregulated normalized enrichment score (ΔNES). A positive ΔNES indicates drug signatures that reverse our COVID-19 signatures, whereas drugs with a negative ΔNES induce signatures similar to the ones observed in COVID-19. Signatures were then grouped by k-means clustering revealing groups of drug signatures that reverse specific patient subgroup signatures (e.g. cluster 5) or those that have the highest impact on all patient subgroups (e.g. cluster 13, **Fig 5c**). Amongst the top signatures in cluster 13 are methylprednisolone (ΔNES_G1_=7.13), immunoglobulins (ΔNES_G1_=6.62), methotrexate (ΔNES_G1_=4.21) and pevonedistat (ΔNES_G1_=4.81) which are all under investigation (clinicaltrials.gov), thereby proving that our *in silico* signature-based drug repurposing approach can indeed predict drugs that have already been deemed potentially beneficial in this disease (**Fig. S7f**). Extracting the leading edge of the most frequently targeted genes by the drugs included in cluster 13 revealed alarmins, such as *S100A8* or *S100A6*, and *SERPINB1*, critical for neutrophil survival by protecting the cell from proteases released into the cytoplasm during stress *(58–60)*. Visualizing these genes in the co-expression network deducted from the blood transcriptomes of our COVID-19 patient cohort identified most of them as part of cluster lightgreen and maroon (**Fig. S7g**). Sample group G1-specific drug signature cluster 5 also encompasses a considerable number of drugs currently being tested in clinical trials to fight COVID-19 (**Fig. 5a+d, Table S9**). Interestingly, a lot of drug signatures in cluster 5 were related to female hormones, such as alpha-estradiol (ΔNES_G1_=2.83), estradiol-cypionate (ΔNES_G1_=2.78), estriol (ΔNES_G1_=2.78), or chlormadinone acetate used in birth control pills (ΔNES_G1_=2.74), but also for example dexamethasone (ΔNES_G1_=2.65) that was recently reported to reduce mortality in severe COVID-19 cases requiring intubation, while showing no benefit for patients with milder disease courses *(61)*. The most frequently targeted genes within the signatures of cluster 5 included protein tyrosine kinase 2 beta *(PTK2B)*, playing an important role for integrin-mediated neutrophil degranulation *(62, 63)*, lysosomal protease Cathepsin D *(CTSD)* expressed in neutrophils and monocytes, as well as the inflammatory mediator Interleukin-1β *(IL1B)* (**Fig. 5e**). The majority of these target genes cluster in the G1-specific lightgreen and pink, as well as in the maroon CoCena^2^ modules. Drugs predicted to be effective for each module are presented as a resource as supplementary information for further inspection (**Table S9**).

We used stratified blood transcriptomes from COVID-19 patients in an *in silico* signature-based approach to identify potential drugs for therapeutic repurposing. Many of our identified hits are indeed already being tested in clinical trials. Further, it became evident that, apart from common therapeutic avenues to address the immune dysregulation in COVID-19 patients, there are patient groups that may benefit from treatments targeting more precisely their immune phenotype and this phenotyping could be used for enrichment of patient groups in clinical trials.

## DISCUSSION

The global spread of SARS-CoV2 resulting in hundreds of thousands of COVID-19 cases urgently demands a more thorough molecular understanding of the pathophysiology of the disease *(12, 17, 64, 65)*. While vaccines are still under development *(66–68)*, therapeutic management of the COVID-19 patients is key to mitigate the clinical burden as well as to prevent deaths. It has become clear that there is great variety in the occurrence of disease manifestation, and dysregulation of local and systemic immune responses have been implicated in disease heterogeneity *(19, 33, 64, 69, 70)*. Here, by applying classical bioinformatics approaches and data-driven co-expression network analysis (CoCena^2^) on blood transcriptomes of COVID-19 patients, we provide evidence for the existence of distinct molecular phenotypes that are not solely explained by current clinical parameters. Particularly in severe COVID-19, we detected dramatic transcriptional changes in the blood compartment with loss of T cell activation and concurrent gain of a rather unique combination of neutrophil activation signals, which was not simply due to changes in cell numbers since isolated neutrophils showed the same transcriptional changes. CoCena^2^ allowed us to group functionally related genes into 10 major transcriptional modules with distinct expression patterns across five, on this basis newly defined COVID-19 patient groups, of which two (G1, G2) were related to severe disease courses. While pronounced neutrophil-related alterations were observed in both subgroups of severe COVID-19 patients (G1, G2), genes associated with coagulation and platelet function were mainly elevated in patients with the most highly elevated number of neutrophils as measured by flow cytometry,an information that was also deduced by linear support vector regression from transcriptome data. Assessment of non-coding RNA species from whole blood transcriptomes also allowed the identification for additional regulatory circuits. For example, we identify *CYTOR*, a lncRNA associated with granulocyte survival *(36)* strongly upregulated in COVID-19 patient group G1, which was accompanied by strong induction of CYTOR interactors such as *VIM* and *PIK3CB (37)*. These findings strongly support the notion that whole blood transcriptomics might not only be suitable for better understanding the systemic immune response in COVID-19 patients, but can also be used to predict novel therapeutic targets involving distinct pathophysiological mechanisms observed in COVID-19. In a ‘reverse transcriptome approach’, we used the specific changes observed in the COVID-19-related transcriptional modules as the bait and searched for inverse correlation in thousands of drug-based transcriptome signatures to predict potential drug candidates. Most interestingly, we identified drug candidates that might be beneficial for all COVID-19 patients, but also candidates that might only be suitable for a subgroup of patients. Lastly, by comparing the transcriptional modules identified in whole blood of COVID-19 patients, we identified unique differences to other viral and bacterial infections, for which similar data were available, suggesting that blood transcriptomes might also be used diagnostically or for outcome prediction in larger clinical cohorts, treatment or vaccine trials in the near future.

Classical bioinformatic assessment of blood transcriptome data comparing defined groups, in this study represented by control individuals and samples derived from either mild or severe COVID-19 patients, already revealed important biology of the systemic immune response. For example, the most significantly elevated transcript was *CD177*, a cell surface molecule on neutrophils, which was enhanced in both mild and severe cases (**Fig. 1, S1**), CD177 has also been introduced as a hallmark for Kawasaki syndrome *(71)*, a syndrome that has been observed in several studies being increased in children and adolescents during the SARS-CoV-2 pandemic *(72–74)*. In acute Kawasaki syndrome, elevated expression of *CD177* was associated with resistance to treatment with intravenous immunoglobulin (IVIG), a therapy in COVID-19 patients that is currently investigated in clinical trials around the world (9 trials, clinicaltrials.gov). Integrating the assessment of CD177 into these trials might help to stratify patients and better predict individual therapy outcome.

Hierarchical clustering of the most variable genes in the dataset already hinted towards further heterogeneity among patients beyond the current clinical differentiation into mild and severe patients (**Fig.1**). Indeed, co-expression network analysis in a data-driven fashion allowed us to define five patient subgroups (G1-5) defined by 10 distinct transcriptional modules, which was corroborated in a second independent cohort (**Fig. 2+S4**). Gene transcription observed in severe COVID-19 patients in G1 clearly differed from severe G2 COVID-19 patients particularly in modules darkgrey, pink, orchid, and maroon (**Fig. 2c**). For example, biological mechanisms related to the darkgrey module included blood coagulation, platelet activation, aggregation and degranulation, as well as cell-cell adhesion and integrin mediated signaling. These are all mechanisms that are integral to several of the complications observed in a subset of severe COVID-19 patients including increased disseminated intravascular coagulation *(75)*, venous thromboembolism *(75, 76)*, stroke *(77)*, or acute cor pulmonale *(78)*, further supporting the need for advanced molecular subtyping of COVID-19 patients, as proposed here based on blood transcriptomes. This is only one prominent example of the rich information within the new structure of molecular COVID-19 phenotypes that we provide here. For further inspection of the data we refer the reader to the online tool that allows to extract module and group specific gene expression information (https://www.fastgenomics.org/).

In addition to many other infectious and non-infectious diseases *(20–28)*, whole blood transcriptomics revealed important insights into the patient structure in COVID-19 and comparative analysis provides first evidence for the unique changes elicited by this disease within the host in comparison to other infections (**Fig. 4**). While cases in G2-4 shared changes with other viral infections such as Chikungunya or Zika, mainly including interferon signature genes *(IFI16, IFI35, IFIT1*, maroon module), partial overlap to bacterial sepsis was observed for G1-G3, albeit the major sepsis module (pink) was not prominently enriched in COVID-19 patients indicating that there are distinct differences in pathology of these two diseases. Although we could establish an integrative model using historical and publicly available blood transcriptome data, we also realized that limited standardization of the experimental procedures (sample processing, library production, sequencing) between different whole blood transcriptomics studies led to the exclusion of several additional important studies. In this context, it will be of great interest whether blood transcriptomics, as it was shown for tuberculosis *(20, 21)*, can be utilized in large enough cohorts and clinical trials for disease risk or outcome prediction in COVID-19. We propose to collect whole blood transcriptomics data in a central registry for direct inspection by the research community and provide a prototype model for such a registry on FASTGenomics. Transcriptome data have been successfully used to predict a role for specific gene networks in the drug response to certain cancer types *(79–83)*. Considering the strong influence of the systemic immune response on severity and outcome of COVID-19, we wanted to establish, whether the global assessment of molecular subgroups of COVID-19 patients could be utilized to predict novel drug targets for this disease addressing the dysregulated peripheral immune response of the host (**Fig. 5**). Using two major databases providing transcriptome signatures to many known drugs, CLUE *(83)* and iLINCS *(82)*, we designed an *in silico* signature-based drug repurposing approach, allowing us to identify candidate drugs *(84)* that might reverse immune pathophysiology as observed in blood transcriptomes. Some of the candidate drugs identified are currently already in clinical trials, for example Imatinib (NCT04394416, NCT04357613, NCT04346147, NCT04356495), Ruxolitinib (NCT04348071, NCT04355793, NCT04377620) or Nintedanib (NCT04338802), for which prediction was particularly high in G1 patients. These trials might benefit from assessing molecular phenotypes of immune cells thereby determining whether patients with G1 type transcriptomes benefit most from such treatment. First study reports have recently declared strong benefit for Dexamethasone treatment in severe COVID-19 cases requiring intubation, while no effect on mortality was seen for those patients who did not require respiratory support *(61)*. Of note, drugs predicted to potentially reverse the transcriptome signatures of the severely affected G1 group may have adverse effects in milder COVID-19 cases from G4 as observed in the contrasting regulation patterns in many of the clusters (**Fig. 5c**). Interestingly and in line with the reports on sexual dimorphism in COVID-19 severity and mortality *(85)*, G1 included only male patients and many of the drugs predicted to reverse the G1-specific signatures were related to female hormones (**Fig. 5d**). However, we also predicted drugs for all COVID-19 patients already in clinical trials such as immunoglobulins (>80 trials, clinicaltrials.gov), or methylprednisolone (>20 trials), findings further supporting the value of our prediction approach. Despite these promising results, strongly suggesting that reverse transcriptomics is not only of value in cancer *(79–81)* but might also be used to identify drugs targeting the immune pathophysiology in COVID-19, we would also like to point out current limitations of our findings that need to be addressed in future studies. Predictions will further benefit from and focused by validation studies in independent COVID-19 patient cohorts, which is to be fostered by a central database for COVID-19 patients’ blood transcriptome data. Nevertheless, we used samples from different countries, illustrating the generalizability. Furthermore, the molecularly derived and prioritized drug candidates presented here might be tested in very recently introduced pre-clinical models *(86)* prior to starting clinical trials. Irrespective of the current shortcomings, we favor such drug candidate identification, since it is based on interrogation of molecular data directly derived from patients’ immune cells involved in the ongoing processes in the disease and therefore may increase the likelihood of a beneficial effect in patients.

Collectively, we provide first evidence for whole blood transcriptomics to potentially become a valuable tool for distinguishing COVID-19 from other infections in cases for which pathogen detection might be difficult, for monitoring and potentially predicting outcome of the disease, to further dissect molecular phenotypes of COVID-19, particularly of the host’s immune system, also along the disease course over time, and to support drug target prediction for subgroups of patients. Clearly, in contrast to more sophisticated higher resolution methods, whole blood transcriptomes can be easily obtained in large clinical cohort studies and large clinical treatment trials yet providing an enormous information content about the molecular reactions of the host’s immune system. We therefore propose a blood transcriptome registry following the model we introduce here on the FASTGenomics platform that would allow the scientific community to utilize the information for new clinical studies and to address further large-scale studies into pathophysiological mechanisms of the disease and enhance the chances of trials to demonstrate a clinical benefit in patients.

## Data Availability

The data that support the findings of this study, including transcriptome data from 60 patients at multiple time points who granted informed consent to share such data, are made available at the European Genome-Phenome Archive (EGA) under accession number EGAS00001004503, which is hosted by the EBI and the CRG.

## Acknowledgements

We thank Claudia Finnemann for perfect technical assistance.

## Deutsche COVID-19 Omics Initiative (DeCOI)

Robert Bals, Alexander Bartholomäus, Anke Becker, Ezio Bonifacio, Peer Bork, Thomas Clavel, Maria Colome-Tatche, Andreas Diefenbach, Alexander Dilthey, Nicole Fischer, Konrad Förstner, Julien Gagneur, Alexander Goesmann, Torsten Hain, Michael Hummel, Stefan Janssen, René Kallies, Birte Kehr, Andreas Keller, Sarah Kim-Hellmuth, Christoph Klein, Oliver Kohlbacher, Jan Korbel, Ingo Kurth, Markus Landthaler, Yang Li, Kerstin Ludwig, Oliwia Makarewicz, Manja Marz, Alice McHardy, Christian Mertes, Markus Nöthen, Peter Nürnberg, Uwe Ohler, Stephan Ossowski, Jörg Overmann, Klaus Pfeffer, Alfred Pühler, Nikolaus Rajewsky, Markus Ralser, Olaf Rieß, Stephan Ripke, Ulisses Nunes da Rocha, Philip Rosenstiel, Antoine-Emmanuel Saliba, Leif Erik Sander, Birgit Sawitzki, Philipp Schiffer, Joachim L. Schultze, Alexander Sczyrba, Oliver Stegle, Jens Stoye, Fabian Theis, Janne Vehreschild, Jörg Vogel, Max von Kleist, Andreas Walker, Jörn Walter, Dagmar Wieczorek, John Ziebuhr

## Author contributions

Conceptualization, ACA, FvdV, MGN, JLS, MK, TU; Methodology, MO, MN-G, LB, NR, KB, RK, TSK, CK, MH, LH, IG, SA, KD, LL, NB, MB, KH, MK, HT, SM, EDD, TU; Formal Analysis, ACA, MO, MN-G, LB, NR, KB, RK, TP, TSK, CK, MH, LH, AH, IG, SA, KD, LL, NB, JG, JS-S, LS, TU; Investigation, ACA, MO, MN-G, LB, NR, KB, RK, TP, TSK, CK, MH, LH, AH, IG, SA, KD, LL, NB, JS-S, JLS, TU; Resources, MM, BK, NK, KG, MS, SD, NR, JR, KMK, MTV, GR, VT, AA, PP, MK, MMBB, JN, AK, EJG-B; Writing – Original Draft, ACA, MO, MN-G, LB, NR, KB, RK, TP, TSK, CK, MH, LH, AH, IG, KD, MvU, AD, JLS, TU; Writing – Review & Editing, ACA, BK, MO, MN-G, LB, NR, KB, RK, AD, FvdV, MGN, JLS, MK, MMBB, JN, EJG-B, TU; Visualization, ACA, MO, MN-G, LB, NR, KB, RK, TP, TSK, CK, MH, LH, AH, IG, SA, KD, MvU, LL, NB, JLS, TU; Supervision, ACA, JLS, MK, MMBB, JN, TU; Funding Acquisition, ACA, JLS, MMBB, EJG-B.

## METHODS

### EXPERIMENTAL MODEL AND SUBJECT DETAILS

#### Human Cohorts

##### Whole blood samples for RNA-seq analysis

The study was conducted between March 13 and March 30, 2020. A total of six ml of blood was sampled from patients with community-acquired pneumonia (CAP) by SARS-CoV-2 within the first 24 hours of hospital admission. CAP was defined as the presence of diffuse infiltrates in chest X-ray or chest computed tomography and positive molecular testing of respiratory secretions for SARS-CoV-2. Exclusion criteria were infection by the human immunodeficiency virus; neutropenia; and any previous intake of immunosuppressive medication (corticosteroids, anti-cytokine biologicals and biological response modifiers). The studies were conducted under the 23/12.08.2019 approval of the Ethics Committee of Sotiria Athens General Hospital; and the 26.02.2019 approval of the Ethics Committee of ATTIKON University General Hospital. Written informed consent was provided by patients or by first-degree relatives in case of patients unable to consent. Patients were classified as severe when they were admitted to the intensive care unit because of need of mechanical ventilation; remaining patients were hospitalized in the ward and were classified as mild. The following information was recorded: white blood cell count and differential; administered treatment; and 28-day outcome. A volume of 2.5 ml of the collected blood was transferred into one PAXgene tube and stored at −80°C. The remaining was used for flow cytometry analysis. A similar amount of blood was sampled from 10 controls fully matched for age, gender and the Charlson’s comorbidity index.

For the second cohort, whole blood samples were collected for RNA-seq analysis in PAXgene tubes from 30 patients upon admission to the Intensive Care Unit of the Radboud university medical centre in Nijmegen, the Netherlands. The study was carried out in accordance with the applicable rules concerning the review of research ethics committees and informed consent. All patients or legal representatives were informed about the study details and could decline to participate. COVID-19 was diagnosed by a positive SARS-CoV-2 RT-PCR test in nasopharyngeal and throat swabs and/or by typical chest CT-scan findings. Exclusion criteria were hematological malignancies and/or active chemotherapy, solid organ transplant, auto-immune diseases, and pre-existent use of high dose corticosteroids.

##### Granulocyte samples for RNA-seq analysis

This study was approved by the Institutional Review board of the University Hospital Bonn (073/19 and 134/20). After providing written informed consent, 11 COVID-19 patients were included in the study. In-patients who were not able to consent at the time of study enrollment, consent was obtained after recovery.COVID-19 patients who tested positive for SARS-CoV-2 RNA in nasopharyngeal swabs were recruited at the Medical Clinic I of the University Hospital Bonn between March 30 and May 17, 2020.

Granulocytes were isolated from EDTA-treated or heparinized peripheral blood by density centrifugation over Pancoll or Ficoll-Paque density centrifugation (density: 1.077g/ml). Granulocyte fractions were then treated with 10ml RBC lysis buffer (Biolegend) for 10min. After RBC lysis, cells were washed with DPBS and recovered by centrifugation at 300xg for 10min. Granulocyte pellets were then lysed with 500µl of QIAzol (QIAGEN), shortly vortexed and incubated 5min at RT prior storage at −80°C until RNA extraction.

##### Rhineland Study as control samples within the integrated dataset for disease comparison

###### Study population

The Rhineland Study is an ongoing community-based cohort study in which all inhabitants of two geographically defined areas in the city of Bonn, Germany aged 30–100 years are being invited to participate. Persons living in these areas are predominantly German with Caucasian ethnicity. Participation in the study is possible by invitation only. The only exclusion criterion is insufficient German language skills to give informed consent.

###### Ethical Approval

Approval to undertake the Rhineland Study was obtained from the ethics committee of the University of Bonn, Medical Faculty. The study is carried out in accordance with the recommendations of the International Conference on Harmonization (ICH) Good Clinical Practice (GCP) standards (ICH-GCP). Written informed consent was obtained from all participants in accordance with the Declaration of Helsinki.

###### Blood withdrawal

Overnight fasting blood was collected from all participants between 7:00 and 9:30 AM, including a PAXgene tube for RNA extraction.

## METHOD DETAILS

### Flow cytometry techniques

Whole blood cells were incubated for 15 minutes in the dark with the monoclonal antibodies anti-CD14 FITC, anti-CD3 FITC, anti-CD4 FITC and anti-CD19 FITC (fluorescein isothiocyanate, emission 525nm,Beckman Coulter); with anti-CD4 PE, anti-CD8 PE, and anti-CD(16+56) PE (phycoerythrin, emission 575nm, Beckman Coulter); and with anti-CD45 PC5 (emission 667nm, Beckman Coulter). Fluorospheres (Beckman Coulter) were used for the determination of absolute counts. Cells were analyzed after running through the CYTOMICS FC500 flow cytometer (Beckman Coulter Co, Miami, Florida). Isotypic IgG controls stained also with anti-CD45 were used for each patient.

### Whole blood RNA isolation

Total RNA was isolated from whole blood samples stored and stabilized in PAXgene RNA tubes using the Qiagen PAXgene Blood miRNA kit according to manufacturer’s guidelines. Eluted RNA was dissolved in RNase free water. The quality and quantity of RNA was evaluated by visualization of 28S and 18S band integrity on a Tapestation 4200 system (Agilent).

### RNA-sequencing

Total RNA was converted into double-stranded cDNA libraries using the TruSeq Stranded Total RNA with Ribo-Zero Globin kit (Illumina). In brief, ribosomal and globin mRNA were depleted from 750ng purified total RNA using biotinylated, target-specific oligos combined with Ribo-Zero rRNA removal beads, remaining RNA was fragmented using divalent cations under elevated temperature. First-strand was generated using SuperScript2 RT (Invitrogen) supplemented with Actinomycin D, followed by second-strand synthesis with dUTP replacing dTTP. 3’ ends were adenylated and index adapters were ligated before subsequent PCR amplification to yield the final library. Remaining overhangs were converted into blunt ends via exonuclease/polymerase activities and enzymes were removed. Selective enrichment of DNA fragments with ligated adaptor molecules was performed using Illumina PCR primers in a 15 cycles PCR reaction, followed by purification cDNA using SPRIBeads (Beckman-Coulter). Libraries were quantified by Qubit dsDNA HS Assay (Thermo Fisher Scientific) and fragment size distribution was determined using the HS D1000 assay on a Tapestation 4200 system (Agilent). High-throughput sequencing was carried out with a NovaSeq™ 6000 Sequencing System S2 (50bp paired-end reads), and data was converted into fastq files using bcl2fastq2 v2.20.

### RNA-sequencing analysis

Sequenced reads were aligned and quantified using STAR: ultrafast universal RNA-seq aligner (v2.7.3a) *(87)* and the human reference genome, GRCh38p13, from the Genome Reference Consortium. Raw counts were imported using DESeqDataSetFromHTSeqCount function from DEseq2 (v1.26.0) *(88)* and rlog transformed according to DEseq2 pipeline. DESeq2 was used for the calculation of normalized counts for each transcript using default parameters. All normalized transcripts with a maximum over all row mean lower than 10 were excluded resulting in 37,526 present transcripts. Differentially expressed genes were calculated for the scenarios status (COVID-19 vs. controls), mild/severe (severe COVID-19 vs controls, mild COVID-19 vs controls, and severe vs mild COVID-19) and new_cluster (1vs6, 2vs6, 3vs6, 4vs6, and 5vs6) separately using a p-value cut-off of 0.05, an adjusted p-value (IHW) < 0.05 (independent hypothesis weighting) and a FC of 2. All present transcripts were used as input for principal component analysis. The top 25% most variable transcripts within the dataset were selected and visualized in a heat map. DEGs were visualized as DE bar plots and were used as input for volcano plots.

### Gene ontology enrichment analysis (GOEA)

To test for functional enrichment within all three scenarios, we performed GOEA for up- or downregulated transcripts in the respective comparison using gene ontology set of biological processes. Gene set “c5.bp.v7.0.symbols.gmt” was obtained from the Molecular Signatures Database (MSigDB) *(89)*. compareCluster and enrichGo functions from the R package ClusterProfiler (v3.12.0) *(90)* were used to determine significant enrichment (q-value<0.05) of biological processes. All present genes were used as background (universe).

### Filtering for transcription factors, epigenome, surfaceome and secretome

All present transcripts were filtered and sorted by their variance in the dataset. The 20 most variable genes of each category were selected and visualized using a heat map. Transcription factor lists were extracted from *(91)*, the epigenome gene list was literature-driven, surface and secretome markers were extracted from the Human Protein Atlas *(92)*.

### Clustering of patients according to clinical parameters

The contribution of each clinical parameter to the transcriptome in COVID-19 patients was determined using linear modelling of each parameter separately with PC1. Clinical parameters with rounded up adjusted r-square ≥0.2 were used for agglomerative hierarchical clustering of the COVID-19 patients. A dissimilarity matrix based on Gower distance was calculated using the daisy function from the cluster packages (version 2.1.0). Agglomerative hierarchical clustering was performed using the hclust function, defining the method with a setting for ward.D2 method linkage. We evaluated the clustering by extracting clusters statistics using the function cluster.stats from the package fpc (version 2.2-5). The number of clusters was chosen at the value at which the lowest distance among patients within clusters (i.e. low value of within-cluster sum of squares distance) and preserving a high distance among clusters (i.e. high average silhouette width) was achieved, while still maintaining a comparable number of individuals among the clusters.

### Linear support vector regression

Linear support vector regression *(38)* was employed to computationally deconvolute the study’s whole blood samples. Gene expression tables were normalized with DESeq2 and were utilized as the input mixture file. LM22-subsetted signatures for B cells, T cells, NK cells, monocytes, dendritic cells, eosinophils and neutrophils were generated as described on https://cibersort.stanford.edu/tutorial.php. The algorithm was subsequently run with 1,000 permutations and the proportions of cell types were visualized with ggplot2 (v3.2.1) *(93)*.

### CoCena^2^: Construction of Co-expression networks, analysis - automated

To define differences and similarities in transcript expression patterns among the different groups, CoCena^2^ (Construction of co-expression Network Analysis – automated) was performed based on Pearson correlation. CoCena^2^ is a network-based approach to identify clusters of genes that are co-expressed in a series of observed conditions based on data retrieved from RNA-sequencing. The tool offers a variety of functions that allow subsequent in-depth analysis of the biological context associated with the found clusters. As input for the analysis the 10,000 most variable genes were used.

To identify genes whose expression patterns are highly similar across all tested samples, pairwise Pearson correlation coefficients are calculated using the R package Hmisc (v4.1-1). The underlying assumption of the Pearson correlation to the data is that it is normally distributed, which is a valid assumption to make in the context of gene expression when looking at expression patterns within different experimental conditions. The correlation between each pair of genes is the basis for the subsequent network construction. Therefore, the tool focuses mainly on positively correlated gene pairs, since the rate of confirmation of an edge representing an association of genes is higher than that of a non-existing association.

In order to refine the structure of the upcoming network and to unravel the condition specific signatures, a correlation cut-off is proposed to mark the minimal correlation a pair of genes must exhibit for their co-expression to be taken into account. The cut-off is determined based on different criteria:

#### 1) Scale-free topology

Gene expression networks have been argued to have a scale-free topology *(94)*, meaning that the majority of vertices has a low number of adjacent edges, also referred to as the vertex’ degree, whereas only very few vertices have a high degree. The degree distribution of scale-free networks asymptotically follows a power law. To assess the scale-free topology of a network constructed by a given correlation cut-off, a log-log plot of the degree distribution is constructed and the R^2^-value of the resulting linear regression is used to evaluate the scale-free criterion.

#### 2) Number of graph components

A graph component is a subset of nodes, such that there is a path from every node within the component to any other node in that same component but none connecting the nodes to any outside of that component. Even though there exist functional collections of genes that cooperate to fulfil a common task, these collections are not expected to be operating independently within the cell. Thus, the cut-off proposal favors graphs with a small number of components.

#### 3) Number of edges

To avoid a highly connected graph with great lack of structure -”hairball”-, the cut-off is chosen such that the number of edges is minimized while respecting the above-mentioned criteria.

A Pearson correlation coefficient cut-off of 0.857 (6,085 nodes and 252,584 edges) was chosen to construct scale-free networks.

The undirected co-expression network is constructed based on the gene pairs which show a higher correlation in their expression pattern than the set cut-off. A series of network-based clustering algorithms is available to then identify clusters of strong co-expression within the network. An option “auto” is provided, which tests the different clustering algorithms and picks the one that achieves the highest modularity score. Unbiased clustering was performed using the “label propagation” algorithm in igraph (v1.2.1) [The igraph software package for complex network research] and was repeated 1,000 times. Genes assigned to more than 5 different clusters during the iterations received no cluster assignment.

To assess the expression strength of the found gene clusters in the different studied conditions, the group fold changes (GFCs) of the conditions are calculated for each gene by calculating the mean expression of a gene over all samples and then computing the fold change of the mean gene expression within each condition from the overall mean. The GFCs of all genes within one cluster are then added and divided by the total number of genes per cluster, resulting in condition-specific GFCs per cluster. Agglomerative hierarchical clustering was performed by the hclust function (cluster package, version 2.1.0), using a dissimilarity matrix of samples based on the GFC values of each sample defined with the daisy function for calculating the Euclidean distances. The number of clusters was set to achieve a low within-cluster sum of squares distance and a high average silhouette, while preserving a comparable number of individuals within each cluster. The clinical parameters and the GFCs results are displayed in a heat map where conditions are clustered by their GFCs revealing similar and opposing patterns (Cluster/Condition heat map).

Utilizing the R-package *clusterProfiler*, CoCena^2^ automatically analyses the gene clusters with respect to different kinds of gene set enrichments: The genes within each cluster are scanned for enrichment in KEGG *(95)*, Hallmark *(96)*, Gene-Ontology terms *(97)* and Reactome *(98)*. Using the R-package *pcaGoPromoter (99)* the genes are also analyzed for enrichment of transcription factor binding sides and if the predicted transcription factors are present in the data, their expression profile is visualized to facilitate evaluation of their possible role.

To investigate the interactions between protein-coding and long-non-coding RNAs, we utilized the enricher function from the clusterProfiler package. We performed an enrichment analysis for lncRNA species, using the protein-coding genes that belong to the lightgreen cluster as the input gene list and all the network protein-coding genes as background. The annotation table defining lncRNA to protein-coding RNA was downloaded from the RNA interactome database RNAInter *(100)*, filtered to only include interactions of lncRNA detected by the RNA sequencing, had an experimental validation score of at least 0.5 and were involved in regulating the function of granulocytes *(36)*. Next, to obtain a comprehensive understanding of the lncRNA that may be relevant for this specific network module, the lncRNA found by the enrichment analysis with p-value <0.1 were sorted according to the highest number of genes. Thereafter, Spearman correlation amongst the gene expression of each lncRNA and its corresponding protein-coding RNAs was performed, and significant protein-coding RNA genes were plotted in a heat map. The CoCena^2^ network was visualized by using the ggplot function from the ggplot2 package. Annotations were generated by filtering the edges of the network for the 5 top connected transcription factors, epigenetic regulators, and surface and secretome markers in each cluster. GO enrichment analysis was performed on each cluster by utilizing the enrichGO function from the clusterProfiler package to assess the overall functionality of the cluster using the genes of each cluster as the input and all the in the network as background. The top GO term and top connected genes of each cluster were compiled representing their general characteristic.

### Granulocyte dataset analysis

Granulocyte raw data was aligned and quantified using STAR (v2.7.3a) and the human reference genome, GRCh38p13, from the Genome Reference Consortium. Raw counts were imported using DESeqDataSetFromHTSeqCount function and rlog transformed. DESeq2 was used for the calculation of normalized counts for each transcript using default parameters. All normalized transcripts with a maximum over all row mean lower than 10 were excluded resulting in 27,323 present transcripts. Differentially expressed genes were calculated for the severe vs mild for day 1-14 and 15-28 post 1st symptoms groups) separately using a p-value cut-off of 0.05, an adjusted p-value (IHW) <0.05 (independent hypothesis weighting) and a FC of 2. All present transcripts were used as input for principal component analysis. DEGs were visualized as DE bar plots.

### Data Integration for Disease Comparison

To describe the differences and similarities between COVID-19 and other diseases, we searched in databases for genomics data such as Gene Expression Omnibus (GEO) *(101)* and ArrayExpress *(102)* [for studies that fulfill certain criteria: I) having at least 20 samples, II) the disease of study was of relevance (other infections, such as bacterial and viral, plus diseases that mainly involve immune dysregulation, such as autoimmune disease) and III) library preparation and sequencing technology differ as little as possible from our COVID-19 protocol. The fastq files of 18 additional studies (588242, GSE101705, GSE107104, GSE112087, GSE127792, GSE128078, GSE129882, GSE133378, GSE143507, GSE57253, GSE63042, GSE66573, GSE79362, GSE84076, GSE89403, GSE90081, GSE97590, GSE99992 and the Rhineland study) were downloaded and aligned with STAR. The counts were imported into R (v3.6.2) and were modelled for each gene using DESeq2. Merged raw counts were filtered for the genes present in the COVID-19 co-expression network, ribosomal protein-coding genes and mitochondrial genes were removed, yielding a total of 5,770 genes and 3,176 samples. To account for differences in sequencing depth across studies, a quantile normalization was performed on the filtered data. Group fold changes were calculated, where the grouping variable was set to be the disease status.

To explore COVID-19 associated expression of genes within the integrated dataset, the data was intersected with the gene modules previously retrieved from the COVID-19 CoCena^2^ network, the mean group-fold-changes were determined per cluster and condition and visualized in a heat map. The modules were analyzed for enriched immune cell markers as provided by CIBERSORT and BD Rhapsody and those that showed neutrophil enrichment were screened for genes representative of different neutrophil subtypes as recently described *(42)*.

### Enrichment of signature from scRNA data of granulocytes

The signatures of different neutrophil states in COVID-19 as previously described *(42)* were enriched for the different clusters from CoCena^2^.

To get a more fine-grained differentiation of the specific neutrophil states for figure 3, the authors kindly provided additional signatures from the scRNA dataset using a Wilcoxon rank sum test for differential gene expression implemented in Seurat. Genes had to be expressed in >10% of the cells of a cluster, exceed a logarithmic threshold >0.1 and to have >5% difference in the minimum detection between two clusters. The following additional comparisons were performed: 8 and 9 (pre- and immature neutrophils combined) VS the rest, 1,3,4,6 (neutrophil states from control patients) VS the rest. To get unique signature genes for clusters 0, 2 and 5 (COVID-19-specific clusters) we took the following approach for each cluster: 1) Calculate DEG for cluster 0 VS all other clusters. 2) Calculate DEG for cluster 0 vs 2&5. 3) Take intersection of these two calculations. 4) Remove genes that occur in more than one of these intersections of cluster 0,2 or 5.

### Gene set enrichment analysis (GSVA)

The GSVA R package (v1.34.0) *(103)* was used to test the enrichment of neutrophil signatures *(42)* in the normalized gene expression table. The *gsva* method was used for the run and data were visualized in a heat map with the pheatmap (v1.0.12) package.

### Overview of drugs

An overview of currently used, recommended or investigated drugs for treatment of COVID-19 patients was compiled from drug lists and lists of drugs in clinical trials downloaded from https://www.drugbank.ca/covid-19, https://www.pharmgkb.org/page/COVID and https://clinicaltrials.gov/ct2/results?cond=COVID-19 (last update: 2020-06-05). Classification of the drugs was performed based on the ATC code, as well as additional research on the drugs action. Drug target genes were identified using the DrugBank database *(104)* (**Table S6**). The number drugs currently recommended or investigated, as well as the number of clinical trials within the respective drug classes were visualized using the ggplot2 package *(105, 106)*. The target genes of the drugs currently recommended or investigated with a minimum frequency of 4 were visualized in a word cloud using the wordcloud package (version 2.6).

### Drug prediction

To identify drugs, which reverse the gene expression signature observed in the comparisons of the COVID-19-specific clusters compared to the control cluster, the drug prediction databases iLINCS (http://www.ilincs.org/ilincs/) and CLUE (https://clue.io/) were accessed. As input for the drug prediction the top 1000 (iLINCS) or the top 100 (CLUE) DEGs were used. Drugs reversing the COVID-19 gene expression signature (defined by a negative score) were pooled together with drugs under investigation in current literature, resulting in a list of 940 unique drugs. Using the iLINCS API (https://github.com/uc-bd2k/ilincsAPI/blob/master/usingIlincsApis.Rmd), every gene expression signature from each drug listed in the signature libraries iLINCS chemical perturbagens (LINCSCP), iLINCS targeted proteomics signatures (LINCSTP), Disease-related signatures (GDS), Connectivity Map signatures (CMAP), DrugMatrix signatures (DM), Transcriptional signatures from EBI Expression Atlas (EBI), Cancer therapeutics response signatures (CTRS) and Pharmacogenomics transcriptional signatures (PG) were downloaded. Labelling was performed in the following principle: “drug name”_”database”_”database ID”. Signatures were ordered by fold change and only the top 300 genes were used. This resulted in a total of 62,897 unique drug signatures each with an up- and down-regulated set. Subsequently, GSEA *(107)* was performed on the sequencing data for every up- and down-regulated set for each drug and each cluster comparison. The resulting normalized enrichment scores (NES) were used to calculate the delta NES for each drug, defined as ΔNES = NES (down) – NES (up), ergo the difference of the NES from the downregulated set and the NES from the upregulated set of each respective drug. These ΔNES values were then k-mean clustered (k=40). The clusters showing the highest ΔNES values for all comparisons and the cluster showing only high ΔNES in the comparison G1vsG6 (most severe) were chosen and selected ones of the uniquely present drugs shown. The leading edge genes of the downregulation signatures of these drugs for theG1vsG6 comparison were examined and the frequency was counted. Recurring target genes were plotted on the CoCena^2^ network.

Patterns of differential gene expression of genes targeted by drugs which are currently approved or under investigation for the treatment of COVID-19 patients were visualized using ggplot2. To this end, target genes of each drug and their first degree neighbors were extracted from several databases and the gene co-expression networks, respectively. Regulation patterns of expression of these genes in different COVID-19 patient groups, as compared to the control group, were classified as up-/downregulated or not significant (n.s.) when pairwise comparisons of gene expression of COVID-19 patients and controls were not statistically significant. The same methodology was applied to genes not included in the drug-target list to identify genes which are not targeted by current drugs but could be potentially targeted by newly identified drugs.

## CONTACT FOR REAGENT AND RESOURCE SHARING

For further information and requests for resources and reagents should be directed to and will be fulfilled by the lead contact, Dr. Thomas Ulas.

## DATA AND SOFTWARE AVAILABILITY

### Data Availability

The data that support the findings of this study, including transcriptome data from 60 patients at multiple time points who granted informed consent to share such data, are made available at the European Genome-Phenome Archive (EGA) under accession number EGAS00001004503, which is hosted by the EBI and the CRG. The Rhineland Study’s dataset is not publicly available because of data protection regulations. Access to data can be provided to scientists in accordance with the Rhineland Study’s Data Use and Access Policy. Requests for further information or to access the Rhineland Study’s dataset should be directed to. In addition to data deposition on EGA, we provide an interactive platform for data inspection and analysis via FASTGenomics (fastgenomics.org). The FASTGenomics platform also provides normalized count tables of the datasets generated in this study. Materials, code, and data are available from the corresponding author upon reasonable request. CoCena^2^ is also available under https://github.com/Ulas-lab/CoCena2.

## FIGURES

**Figure S1 – related to Figure 1.**
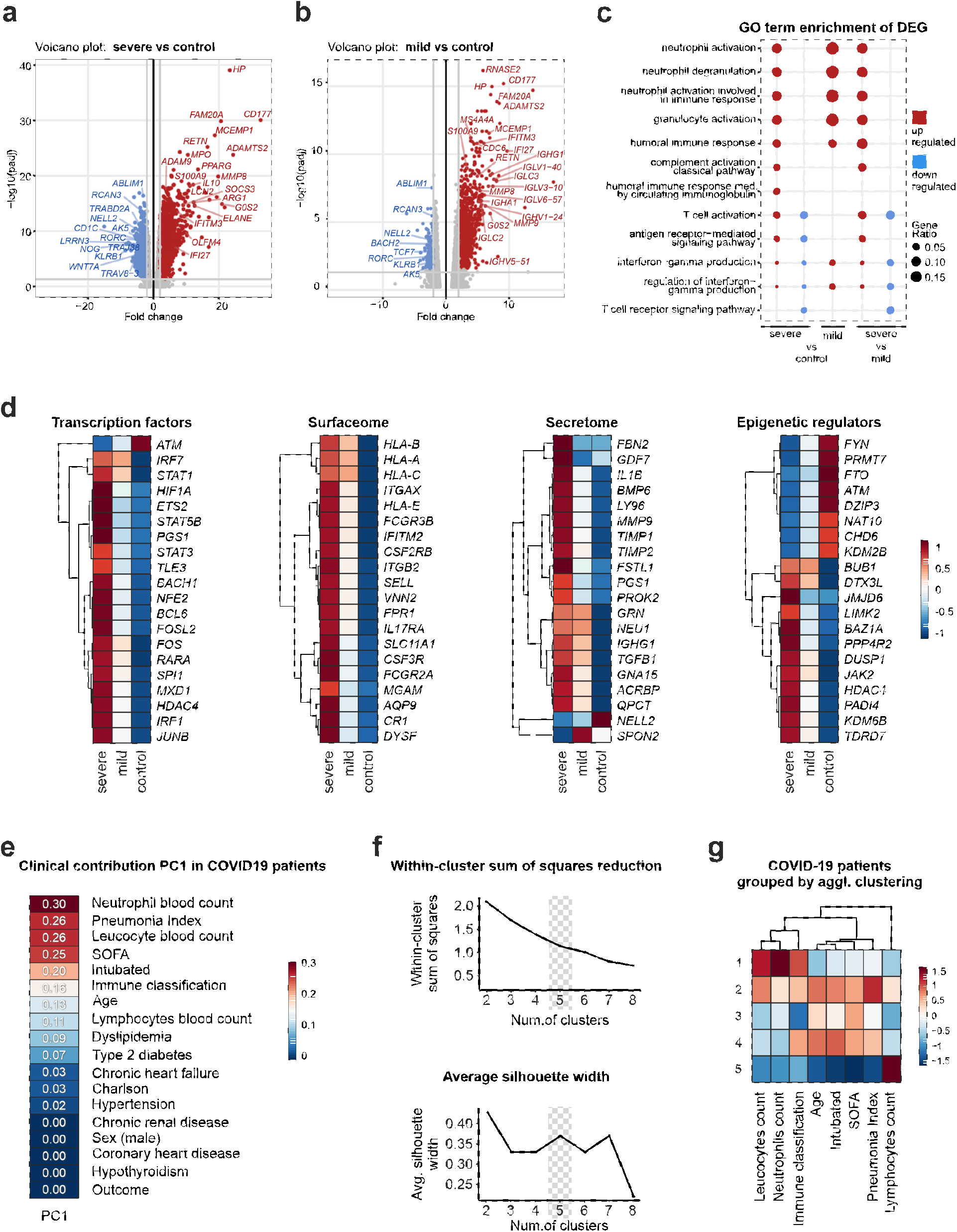
**(a-b)** Volcano plots depicting fold changes and FDR-adjusted p-values comparing severe **(a)** or mild **(b)** COVID-19 patients vs. controls. Differentially expressed up- (red) and downregulated genes (blue) are shown and selected genes are highlighted. **(c)** Plot of top 10 most enriched GO terms for significantly up- and downregulated genes. Ratios of significantly regulated genes within enriched GO terms (GeneRatio) are shown for the comparisons between mild or severe COVID-19 patients and controls as well as between ‘mild’ and ‘severe’ COVID-19 patients. **(d)** Heat map of group mean gene expression values from the top 20 most variant transcription factors, epigenetic regulators, surface and secreted proteins. **(e)** Heat map of the linear model adjusted r-square that includes each clinical parameter with PC1. Clinical parameters with r-adjusted square ≥0.1 were used for agglomerative clustering of COVID-19 patients. **(f)** Plots for agglomerative clustering statistics: within cluster sum of squares and high average silhouette width scores. **(g)** Heat map presenting summary statistics of the clinical parameters used for the clustering across clinical agglomerative clusters 1-5.

**Figure S2 – related to Figure 2.**
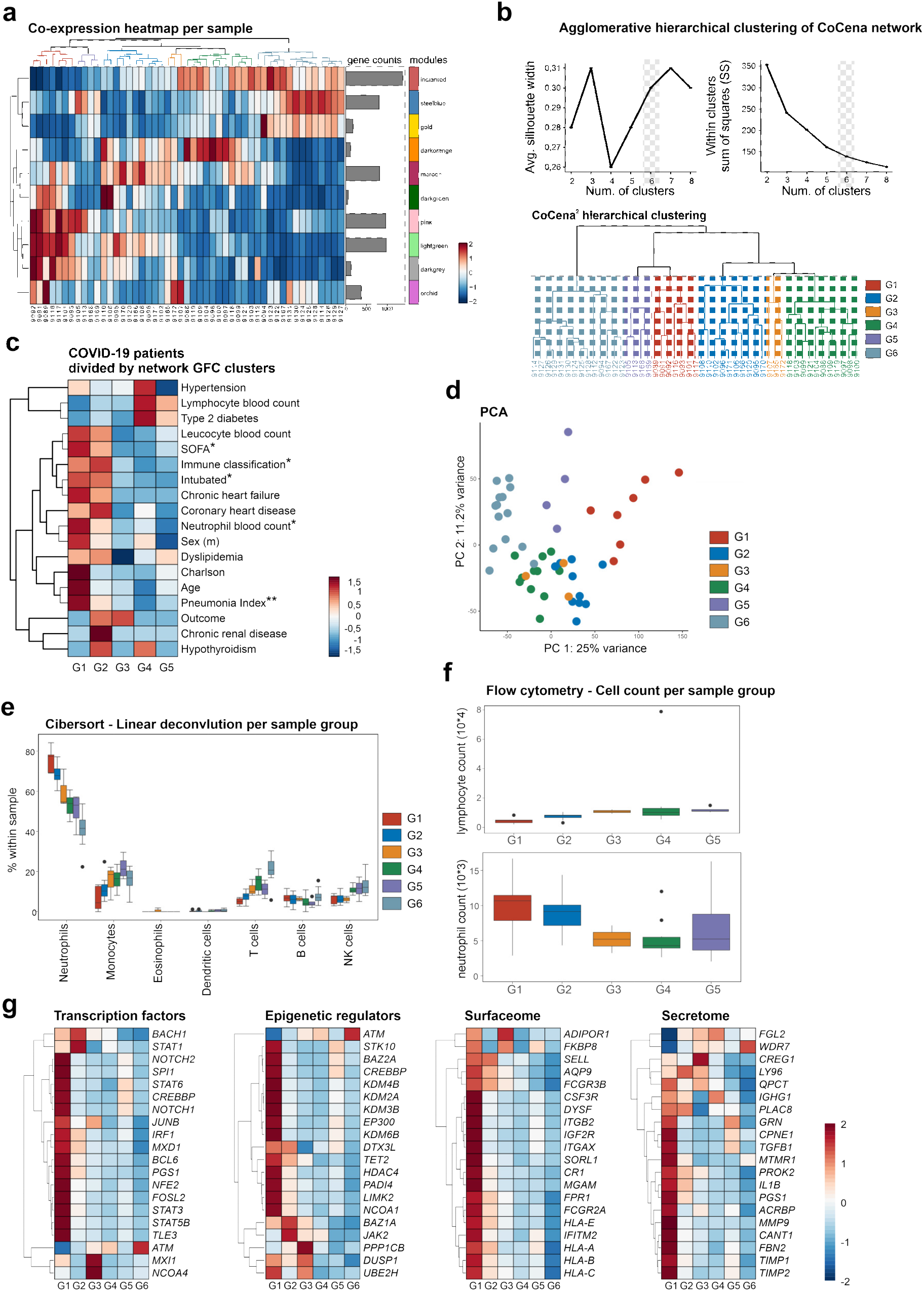
**(a)** Group fold change (GFC) heat map and hierarchical clustering for each sample and the gene modules identified by CoCena^2^ analysis. **(b)** Agglomerative hierarchical clustering of the samples according to the GFC. Top plots present the clustering statistics (within cluster sum of squares and high average silhouette width scores) used for the generation of the six data-driven CoCena^2^ sample groups G1-G6, which are plotted in the dendrogram plot. **(c)** Heat map presenting summary statistics of clinical parameters for COVID-19 patients grouped according to the CoCena^2^ sample groups G1-G5 Presented are scaled values of the mean value of the parameters age/blood cell counts/SOFA score/Pneumonia Index/Charlson score, and prevalence of the comorbidity/death (outcome)/male (sex)/immune classification (intermediate/dysregulation/MAS). Statistical differences were estimated among the groups via the one-sided Anova test or Fisher test, for numeric or categorical values respectively. (*,** p-value < 0.05, 0.01 respectively) **(d)** PCA plot depicting relationship of all samples based on dynamic gene expression of all genes. Coloring based on the six data-driven CoCena^2^ sample groups G1-G6. **(e)** Cibersort cell type deconvolution at cell subset level. Grouping based on the six data-driven CoCena^2^ sample groups G1-G6. **(f)** Flow cytometry analysis, number of lymphocytes (upper) and neutrophils (lower) per µl of blood. Grouping based on the six data-driven CoCena^2^ sample groups G1-G6. **(g)** Heat map of DE and top 20 most variable transcription factors, epigenetic regulators, surface and & secreted proteins.

**Figure S3 – related to Figure 2.**
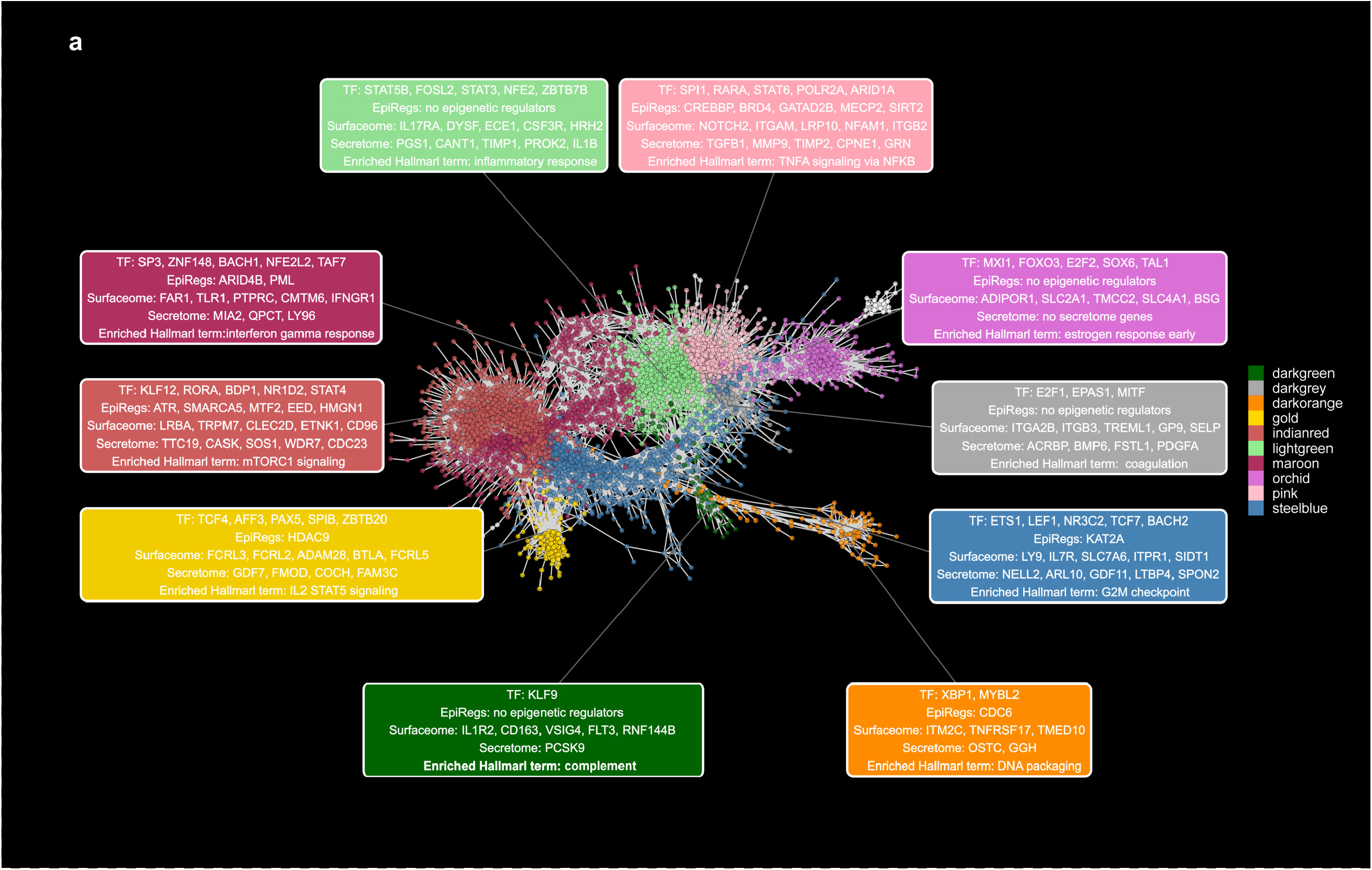
**(a)** Visualization of the COVID-19 CoCena^2^ network. Nodes are genes and edges represent co-expressed genes. Additional module information is displayed by module-colored labels. Labels include information about top-connected transcription factors (TFs), epigenetic regulators, surface & secreted markers as well as representative Hallmark terms.

**Figure S4 – related to Figure 2.**
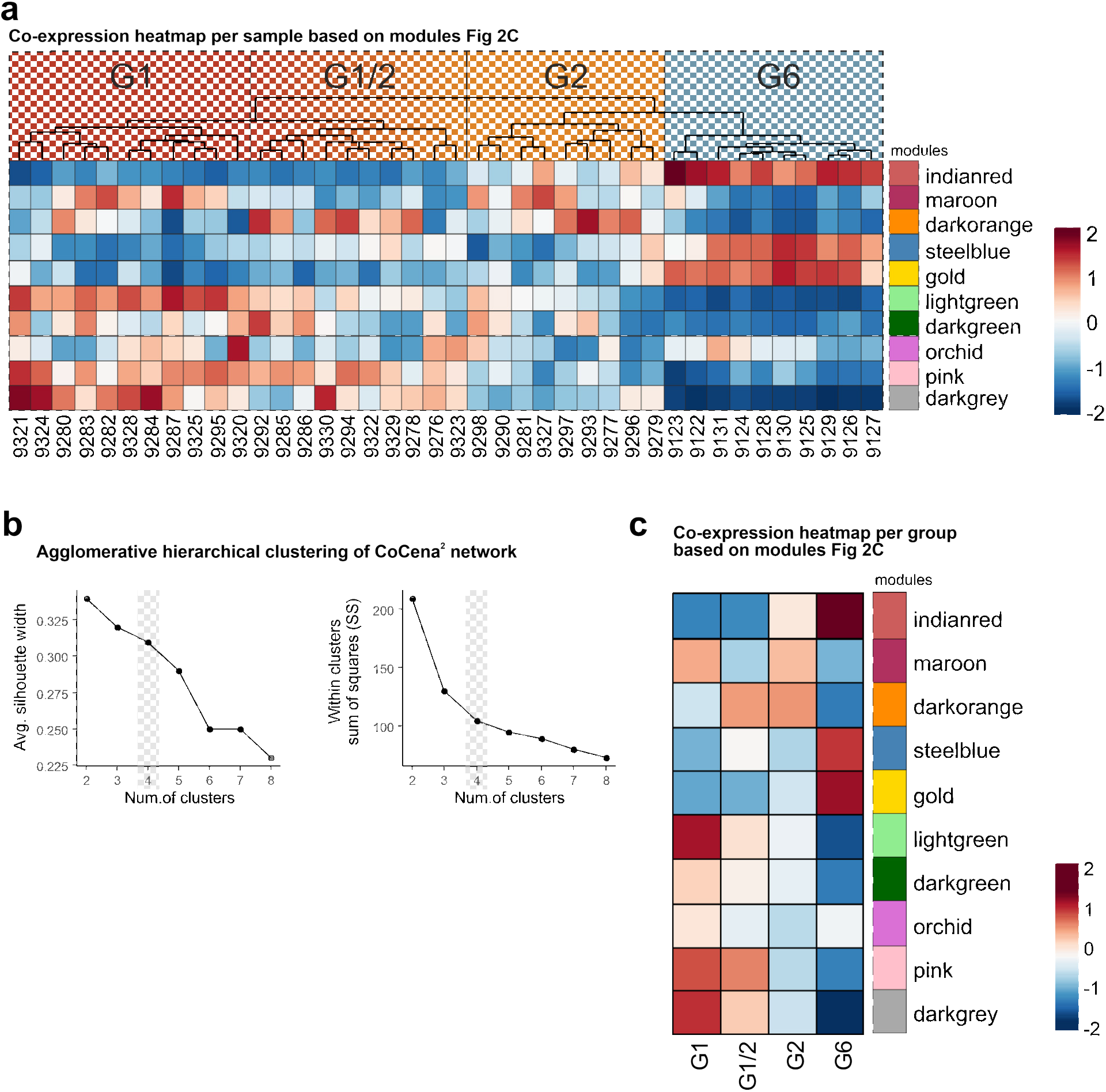
**(a)** Heat map of mean group fold changes (GFCs) of the CoCena^2^ whole blood modules in the second COVID-19 cohort for each sample. Patients are clusters by the mean GFC module expression. Severity patterns found in the whole blood CoCena^2^ network were identified and patients groups were labeled accordingly (G1-G6). **(b)** Agglomerative hierarchical clustering of the samples according to the GFC. Top plots present the clustering statistics (within cluster sum of squares and high average silhouette width scores) used for the generation of the six data-driven CoCena^2^ sample groups G1-G6, which are plotted in the dendrogram plot. **(c)** GFC heat map and hierarchical clustering for the four identified sample groups and the gene modules identified with CoCena^2^ analysis.

**Figure S5 – related to Figure 3.**
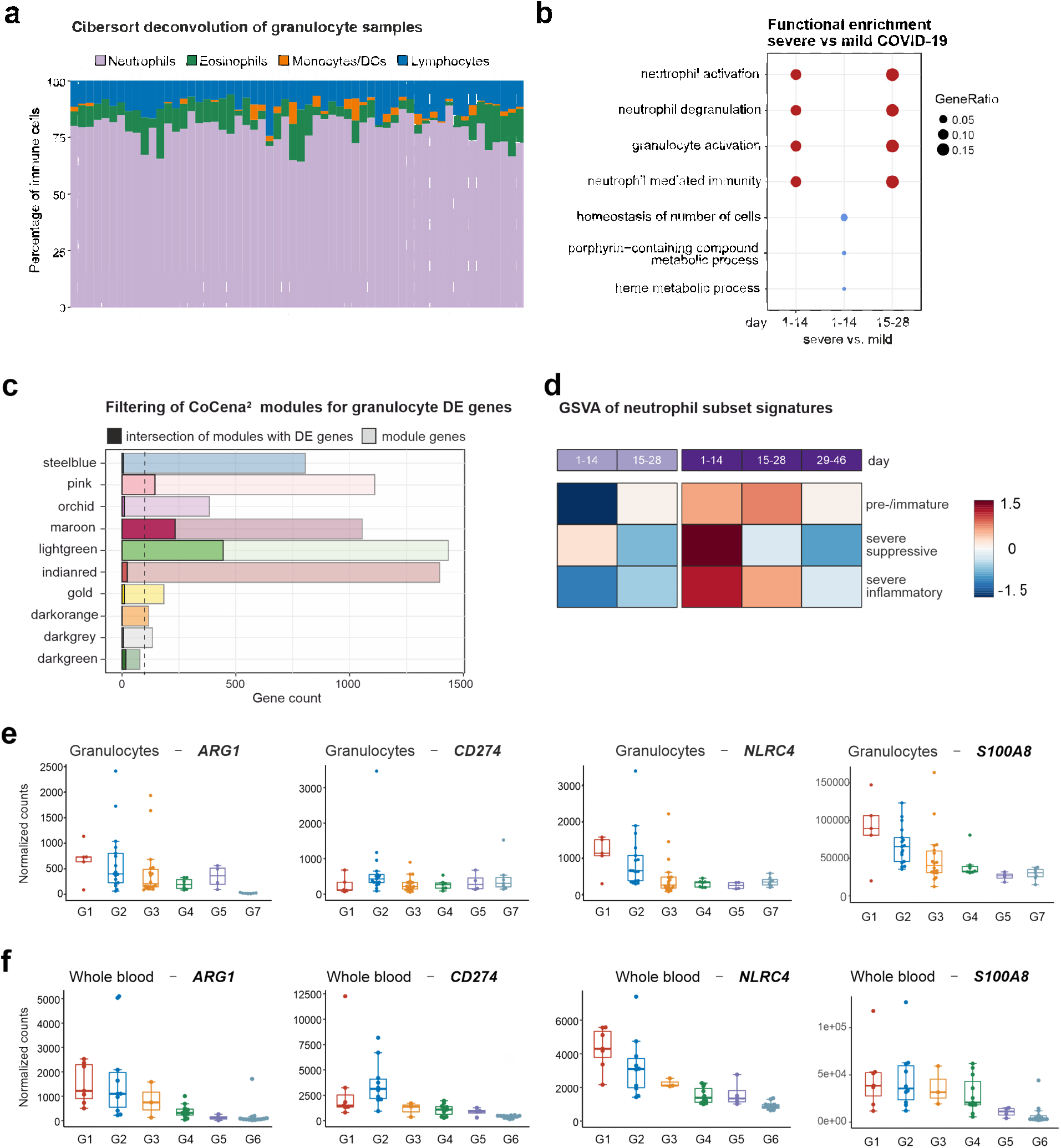
**(a)** Cibersort computational deconvolution of the 59 granulocyte samples used in Figure 3. The LM22 reference signature was used. **(b)** Functional enrichment analysis of the DEGs between severe and mild COVID-19 patients by GOEA. **(c)** CoCena^2^ whole blood module genes filtered for severe vs mild COVID-19 DEGs. Intersection of modules with DEGs are shown in opaque bars, filtered network module genes are shown in transparent bars. **(d)** GSVA of single-cell neutrophil signatures *(42)*. Samples are ordered by COVID-19 severity status and days after disease onset. **(e)** Box plots of *ARG1, CD274, NLRC4* and *S100A6* in granulocytes grouped by G1-G7. **(f)** Box plot of *ARG1, CD274, NLRC4* and *S100A6* in whole blood grouped by G1-G6.

**Figure S6 – related to Figure 4.**
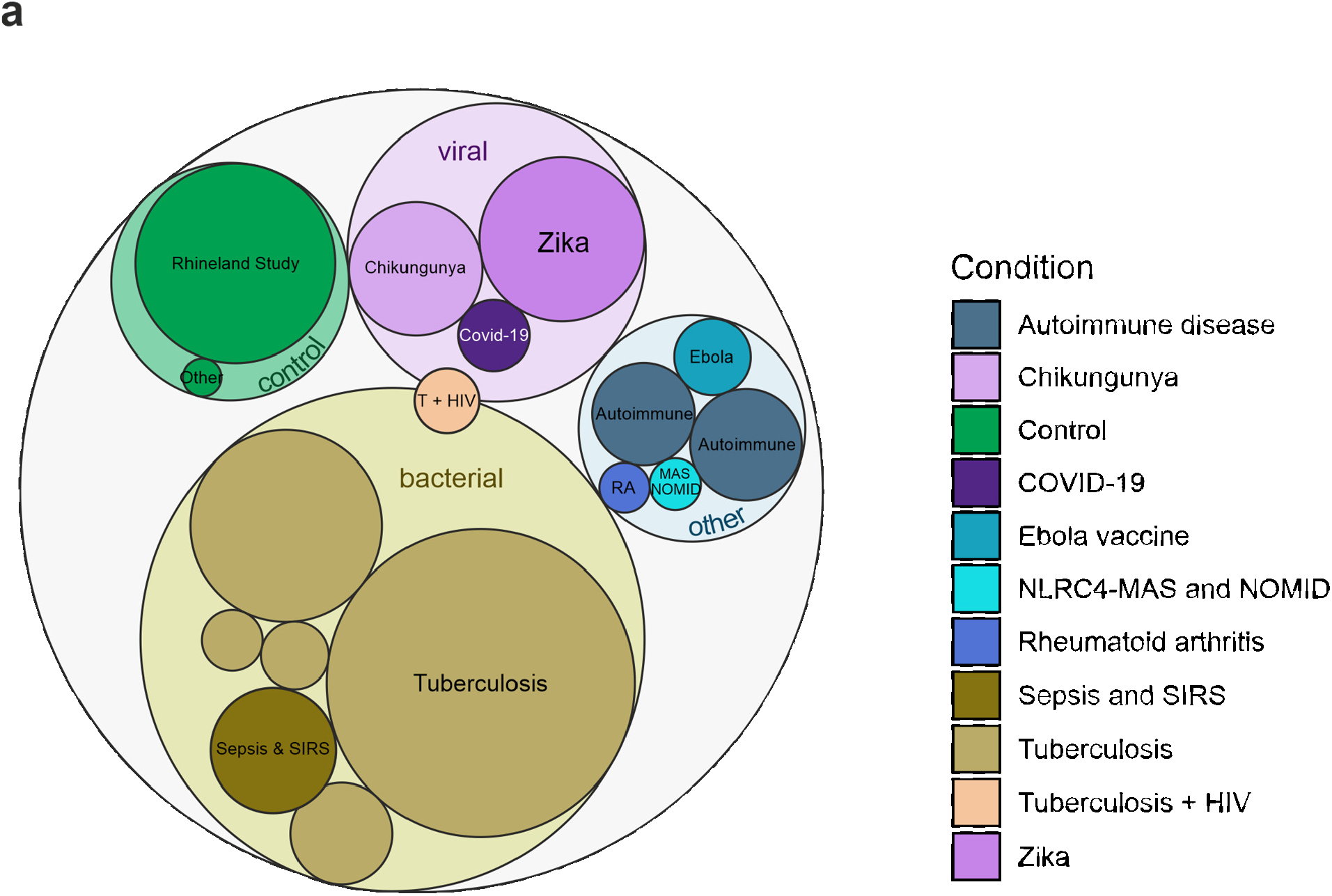
**(a)** Overview of the composition of the integrated dataset comprising 2,817 samples: 39 COIVD-19 samples and 2,778 other conditions and controls.

**Figure S7 – related to Figure 5.**
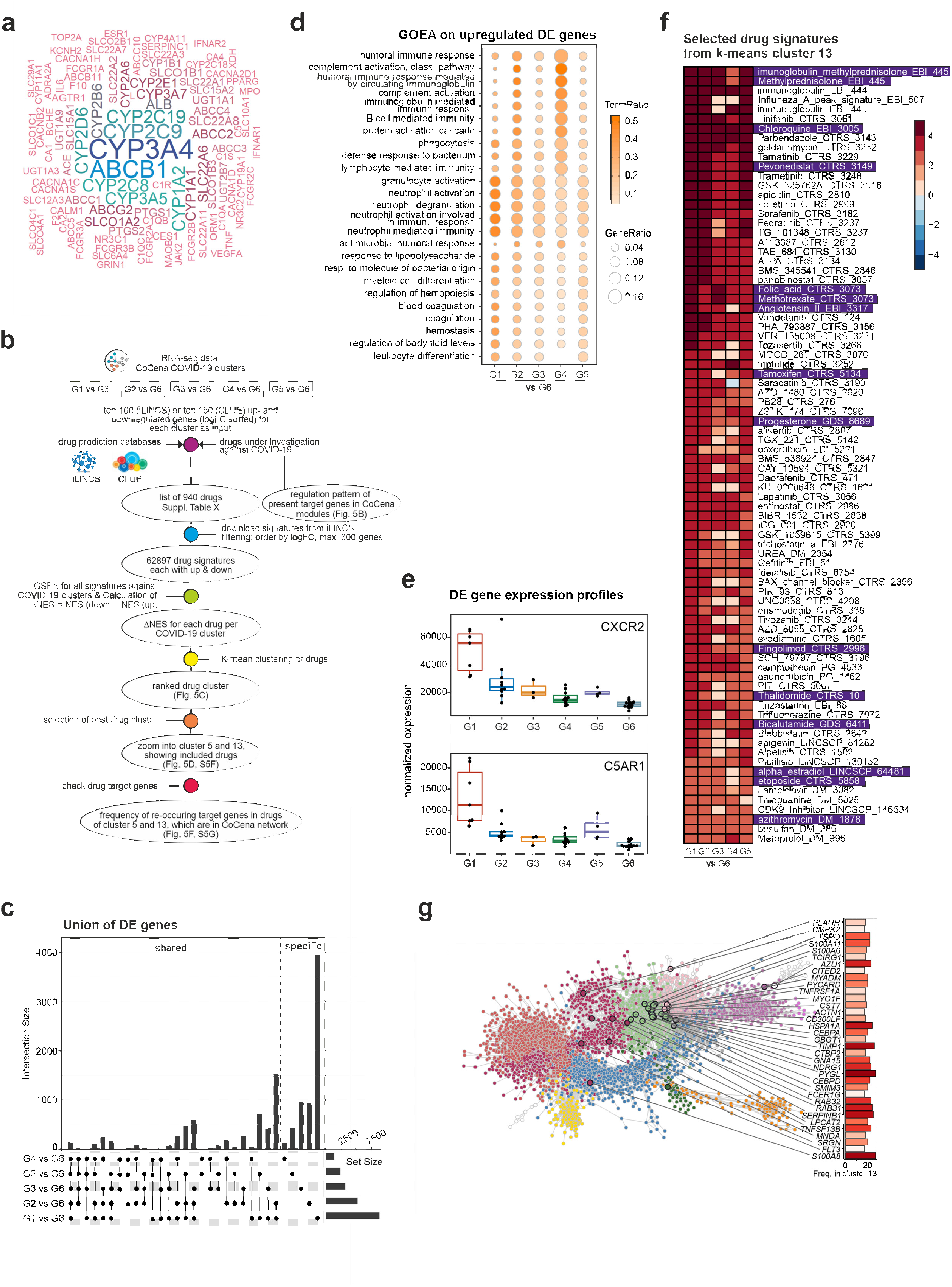
**(a)** Word cloud of the target genes of drugs currently investigated for the treatment of COVID-19 patients. Increasing frequency is represented by increasing size with min. frequency = 41 and max. frequency = 97. **(b)** Schematic workflow of the drug prediction analysis. Drug signatures were collected using the platforms iLINCS and CLUE. Signatures were selected by highest counteracting ΔNES score and combined with signatures of drugs under investigation from the literature. **(c)** Differentially expressed genes (FC>|2|, FDR-adj. p-value <0.05) of comparisons between groups G1-5 vs G6. Vertical bar plots indicate the number of group-specific differentially expressed genes (DEGs, right) and genes shared by several groups (left), whereby the contributing groups are indicated as connected dots (bottom). Horizontal bar plots visualize the size of DEGs per group. **(d)** Gene ontology enrichment analysis of upregulated DEGs obtained for each comparison G1-G5 vs G6. Visualized are significant enrichments (adj. p-value<0.05, q-value<0.05) for the union of top 10 terms per comparison. Term ratio indicates the ratio of DEGs matching the term and the total gene number of that term. **(e)** Box plots of normalized expression of selected DEGs, upregulated in at least one comparison. **(f)** Display of selected drug signatures from k-means cluster 13 from Fig. 5c showing high ΔNES scores throughout all patient groups compared to G6. **(g)** Visualization of recurring target genes in the G1 vs G6 comparison of cluster 13 signatures and their frequency mapped onto the CoCena^2^ network

## REFERENCES

1. N. D. Grubaugh, M. E. Petrone, E. C. Holmes, We shouldn’t worry when a virus mutates during disease outbreaks. Nat. Microbiol. 5 (2020), pp. 529–530.

2. J. R. Fauver et al., Cell. 181 (2020), doi:10.1016/j.cell.2020.04.021.

3. P. Zhou et al., Nature. 579, 270–273 (2020).

4. C. Brignola et al., J. Clin. Gastroenterol. 10, 631–634 (1988).

5. W. Guan et al., N. Engl. J. Med. 382, 1708–1720 (2020).

6. C. Huang et al., Lancet. 395, 497–506 (2020).

7. F. Zhou et al., Lancet. 395, 1054–1062 (2020).

8. D. Wang et al., JAMA - J. Am. Med. Assoc. 323, 1061–1069 (2020).

9. E. Z. Ong et al., Cell Host Microbe. 27 (2020), doi:10.1016/j.chom.2020.03.021.

10. B. Wang, R. Li, Z. Lu, Y. Huang, Aging (Albany. NY). 12, 6049–6057 (2020).

11. W. J. Guan et al., Eur. Respir. J. 55 (2020), doi:10.1183/13993003.00547-2020.

12. R. T. Gandhi, J. B. Lynch, C. del Rio, N. Engl. J. Med. (2020), doi:10.1056/nejmcp2009249.

13. S. A. Dugger, A. Platt, D. B. Goldstein, Drug development in the era of precision medicine. Nat. Rev. Drug Discov. 17 (2018), pp. 183–196.

14. A. Zumla et al., Towards host-directed therapies for tuberculosis. Nat. Rev. Drug Discov. 14 (2015), pp. 511–512.

15. A. Mullard, Coordinating the COVID-19 pipeline. Nat. Rev. Drug Discov. 19 (2020), p. 298.

16. H. Ledford, Nature. 581 (2020), doi:10.1038/d41586-020-01367-9.

17. D. A. Berlin, R. M. Gulick, F. J. Martinez, N. Engl. J. Med. (2020), doi:10.1056/nejmcp2009575.

18. G. Dimopoulos et al., Cell Host Microbe (2020), doi:10.1016/j.chom.2020.05.007.

19. Y. Jamilloux et al., Should we stimulate or suppress immune responses in COVID-19? Cytokine and anti-cytokine interventions. Autoimmun. Rev. 19 (2020),, doi:10.1016/j.autrev.2020.102567.

20. D. E. Zak et al., Lancet. 387, 2312–2322 (2016).

21. E. G. Thompson et al., Tuberculosis. 107, 48–58 (2017).

22. S. Leong et al., Tuberculosis. 109, 41–51 (2018).

23. S. Verma et al., BMC Infect. Dis. 18 (2018), doi:10.1186/s12879-018-3127-4.

24. E. L. Tsalik et al., Genome Med. 6 (2014), doi:10.1186/s13073-014-0111-5.

25. A. Rechtien et al., Cell Rep. 20, 2251–2261 (2017).

26. D. Michlmayr et al., Mol. Syst. Biol. 14 (2018), doi:10.15252/msb.20177862.

27. J. A. Hill et al., J. Virol. 93 (2018), doi:10.1128/jvi.01419-18.

28. E. Bartholomeus et al., J. Transl. Med. 17 (2019), doi:10.1186/s12967-019-2037-6.

29. X. Yang et al., Lancet Respir. Med. 8, 475–481 (2020).

30. G. Chen et al., J. Clin. Invest. 130, 2620–2629 (2020).

31. E. J. Giamarellos-Bourboulis et al., Cell Host Microbe (2020), doi:10.1016/j.chom.2020.04.009.

32. J. Hadjadj et al., medRxiv, in press, doi:10.1101/2020.04.19.20068015.

33. M. Merad, J. C. Martin, Pathological inflammation in patients with COVID-19: a key role for monocytes and macrophages. Nat. Rev. Immunol. 20 (2020),, doi:10.1038/s41577-020-0331-4.

34. P. Lalezari, G. B. Murphy, F. H. Allen, J. Clin. Invest. 50, 1108–1115 (1971).

35. R. Grieshaber-Bouyer, P. A. Nigrovic, Neutrophil heterogeneity as therapeutic opportunity in immune-mediated disease. Front. Immunol. 10 (2019),, doi:10.3389/fimmu.2019.00346.

36. X. Tian, J. Tian, X. Tang, J. Ma, S. Wang, J. Hematol. Oncol. 9 (2016), doi:10.1186/s13045-016-0333-7.

37. Y. Zhang et al., Biomed. Pharmacother. 94, 644–651 (2017).

38. A. M. Newman et al., Nat. Methods. 12, 453–457 (2015).

39. W. Shang et al., J. Med. Virol. (2020), doi:10.1002/jmv.26031.

40. X. Yan et al., J. Med. Virol. (2020), doi:10.1002/jmv.26061.

41. C. Wenham, J. Smith, R. Morgan, COVID-19: the gendered impacts of the outbreak. Lancet. 395 (2020), pp. 846–848.

42. J. Schulte-Schrepping et al., medRxiv, in press, doi:10.1101/2020.06.03.20119818

43. N. Romberg et al., Nat. Genet. 46, 1135–1139 (2014).

44. S. W. Canna et al., Nat. Genet. 46, 1140–1146 (2014).

45. D. Michlmayr et al., Cell Rep. 31 (2020), doi:10.1016/j.celrep.2020.107569.

46. L. S. de Araujo et al., Front. Microbiol. 7 (2016), doi:10.3389/fmicb.2016.01586.

47. W. A. Figgett et al., Clin. Transl. Immunol. 8 (2019), doi:10.1002/cti2.1093.

48. K. Shchetynsky et al., Arthritis Res. Ther. 19 (2017), doi:10.1186/s13075-017-1220-5.

49. R. D. Gray et al., J. Inflamm. (United Kingdom). 10 (2013), doi:10.1186/1476-9255-10-12.

50. A. S. Rohrbach, D. J. Slade, P. R. Thompson, K. A. Mowen, Activation of PAD4 in NET formation. Front. Immunol. 3 (2012),, doi:10.3389/fimmu.2012.00360.

51. A. Carestia et al., J. Leukoc. Biol. 99, 153–162 (2016).

52. N. L. Denning, M. Aziz, S. D. Gurien, P. Wang, Damps and nets in sepsis. Front. Immunol. 10 (2019),, doi:10.3389/fimmu.2019.02536.

53. B. McDonald et al., Blood. 129, 1357–1367 (2017).

54. P. Jha, H. Das, Int. J. Mol. Sci. 18 (2017), doi:10.3390/ijms18112383.

55. T. Németh, M. Sperandio, A. Mócsai, Neutrophils as emerging therapeutic targets. Nat. Rev. Drug Discov. 19 (2020), pp. 253–275.

56. M. Pilarczyk et al., bioRxiv, 826271 (2019).

57. S. M. Corsello et al., The Drug Repurposing Hub: A next-generation drug library and information resource. Nat. Med. 23 (2017), pp. 405–408.

58. M. Baumann, C. T. N. Pham, C. Benarafa, Blood. 121, 3900–3907 (2013).

59. A. Torriglia, E. Martin, I. Jaadane, The hidden side of SERPINB1/Leukocyte Elastase Inhibitor. Semin. Cell Dev. Biol. 62 (2017), pp. 178–186.

60. L. Duplomb et al., J. Mol. Med. 97, 633–645 (2019).

61. “Low-cost dexamethasone reduces death by up to one third in hospitalised patients with severe respiratory complications of COVID-19” (2020), (available at https://www.recoverytrial.net/files/recovery_dexamethasone_statement_160620_v2final.pdf).

62. M. Fuortes, M. Melchior, H. Han, G. J. Lyon, C. Nathan, J. Clin. Invest. 104, 327–335 (1999).

63. L. A. Kamen, J. Schlessinger, C. A. Lowell, J. Immunol. 186, 1656–1665 (2011).

64. W. C. Koff, M. A. Williams, N. Engl. J. Med. (2020), doi:10.1056/nejmp2006761.

65. L. Bao et al., Nature (2020), doi:10.1038/s41586-020-2312-y.

66. N. Lee, A. McGeer, The starting line for COVID-19 vaccine development. Lancet. 395 (2020), pp. 1815–1816.

67. N. Lurie, M. Saville, R. Hatchett, J. Halton, Developing covid-19 vaccines at pandemic speed. N. Engl. J. Med. 382 (2020), pp. 1969–1973.

68. E. Callaway, Nature. 580, 576–577 (2020).

69. K. Subbarao, S. Mahanty, Immunity. 52, 905–909 (2020).

70. N. Vabret et al., Immunity (2020), doi:10.1016/j.immuni.2020.05.002.

71. Y. H. Huang, M. H. Lo, X. Y. Cai, S. F. Liu, H. C. Kuo, Pediatr. Rheumatol. 17 (2019), doi:10.1186/s12969-019-0315-8.

72. J. Toubiana et al., BMJ. 369, m2094 (2020).

73. R. M. Viner, E. Whittaker, Kawasaki-like disease: emerging complication during the COVID-19 pandemic. Lancet. 395 (2020),, doi:10.1016/S0140-6736(20)31129-6.

74. G. Ronconi et al., J. Biol. Regul. Homeost. Agents. 34 (2020), doi:10.23812/EDITORIAL-RONCONI-E-59.

75. H. Al-Samkari et al., Blood (2020), doi:10.1182/blood.2020006520.

76. F. A. Klok et al., Thromb. Res. 191 (2020), doi:10.1016/j.thromres.2020.04.013.

77. T. J. Oxley et al., N. Engl. J. Med. 382, e60 (2020).

78. C. Creel-Bulos et al., Acute cor pulmonale in critically ill patients with covid-19. N. Engl. J. Med. 382 (2020),, doi:10.1056/NEJMc2010459.

79. J. W. Tyner et al., Nature. 562, 526–531 (2018).

80. F. Rambow et al., Cell. 174, 843-855.e19 (2018).

81. Z. Qiu et al., Cancer Cell. 36, 179-193.e11 (2019).

82. A. B. Keenan et al., The Library of Integrated Network-Based Cellular Signatures NIH Program: System-Level Cataloging of Human Cells Response to Perturbations. Cell Syst. 6 (2018), pp. 13– 24.

83. A. Subramanian et al., Cell. 171, 1437-1452.e17 (2017).

84. Y. Zhou et al., Cell Discov. 6 (2020), doi:10.1038/s41421-020-0153-3.

85. E. P. Scully, J. Haverfield, R. L. Ursin, C. Tannenbaum, S. L. Klein, Considering how biological sex impacts immune responses and COVID-19 outcomes. Nat. Rev. Immunol. (2020),, doi:10.1038/s41577-020-0348-8.

86. J. Shi et al., Science. 368, 1016–1020 (2020).

87. A. Dobin et al., STAR: Ultrafast universal RNA-seq aligner. Bioinformatics. 29 (2013), pp. 15–21.

88. M. I. Love, W. Huber, S. Anders, Genome Biol. 15 (2014), doi:10.1186/s13059-014-0550-8.

89. A. Liberzon et al., Bioinformatics. 27, 1739–40 (2011).

90. G. Yu, L. G. Wang, Y. Han, Q. Y. He, Omi. A J. Integr. Biol. 16, 284–287 (2012).

91. D. L. Fulton et al., Genome Biol. 10 (2009), doi:10.1186/gb-2009-10-3-r29.

92. M. Uhlén et al., Science (80-.). 347 (2015), doi:10.1126/science.1260419.

93. K. Ito, D. Murphy, CPT Pharmacometrics Syst. Pharmacol. 2 (2013), doi:10.1038/psp.2013.56.

94. P. Aloy, R. B. Russell, EMBO Rep. 5, 349–350 (2004).

95. M. Kanehisa, S. Goto, Nucleic Acids Res. 28, 27–30 (2000).

96. A. Liberzon et al., Cell Syst. 1, 417–425 (2015).

97. Gene Ontology Consortium, Nucleic Acids Res. 43, D1049–56 (2015).

98. A. Fabregat et al., Nucleic Acids Res. 46, D649–D655 (2018).

99. M. Hansen et al., PLoS One. 7 (2012), doi:10.1371/journal.pone.0032394.

100. Y. Lin et al., Nucleic Acids Res. 48, D189–D197 (2020).

101. E. Clough, T. Barrett, in Methods in Molecular Biology (Humana Press Inc., 2016), vol. 1418, pp. 93–110.

102. A. Athar et al., Nucleic Acids Res. 47, D711–D715 (2019).

103. S. Hänzelmann, R. Castelo, J. Guinney, BMC Bioinformatics. 14 (2013), doi:10.1186/1471-2105-14-7.

104. D. S. Wishart et al., Nucleic Acids Res. 46, D1074–D1082 (2018).

105. H. Wickham, Ggplot2?: elegant graphics for data analysis (Springer, 2009).

106. N. Neth, H.; Gradwohl, unikn: Graphical elements of the University of Konstanz’s corporate design. Soc. Psychol. Decis. Sci. (2019), (available at https://www.spds.uni-konstanz.de/publication-page/unikn-graphical-elements-university-konstanzs-corporate-design).

107. G. Korotkevich, V. Sukhov, A. Sergushichev, bioRxiv, 060012 (2019).

